# Searching for What You Can’t See - Evaluation of Pesticide Residues in Grain Sold at Selected Markets of Southwest Nigeria

**DOI:** 10.1101/2022.12.09.22283068

**Authors:** Modupe Abeke Oshatunberu, Adebayo Oladimeji, Sawyerr Olawale Henry, Morufu Olalekan Raimi

## Abstract

**Background:** Studies on the use of pesticides in southwest Nigeria have revealed a substantial rise in a variety of pesticide-related illnesses, including mental impairment and reproductive problems. Those who work in agriculture and are regularly exposed to pesticides are the most impacted. The World Health Organization (WHO) predicts that three million severe pesticide poisoning episodes occur globally each year, with at least 300.000 deaths and 99% of cases occurring in low- and middle-income nations. The effects of longer-term exposure to pesticides on health are not yet precisely estimated.

**Objectives:** To this end, the objective of this study is to assess the knowledge of pesticide residues and common pesticides in grain-based food (brown and white beans, yellow and white maize, brown millet and rice) about pesticide use in selected markets of Southwest Nigeria. The markets were Oja-titun (market) Ile-Ife, Osun; Alesinloye Market Ibadan, Oyo; Oja Oba, Ado-Ekiti, Ekiti; and Oja Oba, Akure Ondo State. The identification and quantification of pesticide residues was done using a Varian 3800/4000 gas chromatograph mass spectrometer while 60 structured questionnaires were administered to respondents including food merchants, buyers/consumers as well as food vendors.

**Methods:** A total of 240 respondents were selected from four states (Ado-Ekiti, Ibadan, Osun and Ondo) with the aid of structured questionnaire and interview guide using purposive sampling technique. Data were analysed using descriptive statistics.

**Results:** Up to 50.4% never read instructions on pesticide application while a shared 24.8% read them always and occasionally. The number of respondents who never read the instructions were particularly high in Ekiti and Ondo, up to 52.5% of the respondents in Ekiti do not know whether or not they are exposed to pesticides during application. Also, majority of the respondents never attended any professional training on pesticide application, this amounted to 79.3%. Most people were not aware that unsafe pesticide application is harmful to human health. Similarly, a wide variety of symptoms were reported by respondents following pesticide application or consumption of grains, these included headaches and dizziness, itching and redness of the eyes, skin allergy, diarrhea, and stomach disorder, vomiting and loss of appetite, weakness of the body, asthma, permanent skin patches, shortness of breath, excessive sweating. Millet recorded the highest number of OCP residues while maize had the lowest. On the contrary, maize had the highest number of OPP and carbamate residues while millet had the lowest number of OPP residues. Meanwhile, maize, rice and beans had only one carbamate pesticide residue.

**Conclusions:** According to the study’s conclusions, farmers who can only get information from agricultural extension officials should receive urgent and immediate attention for raising awareness. Additionally, extensive IPM training programs must be created with the intention of disseminating precautions for protecting human health and a healthy agro-ecosystem. In order to find more effective pest management methods that utilize less pesticides, it is crucial to reevaluate the pesticide residues and common pesticides found in grains in the targeted markets. To reduce farmers’ exposure to pesticides, it is also required to establish personal protective measures, special educational initiatives, and legislation promoting the use of safer pesticides.

**Significance and novelty:** This study gives policymakers a comprehensive understanding of the ways that may be utilized to close the significant knowledge gap on pesticide residues in grains and provides an insight into the knowledge of pesticide residues and common pesticides in grains.

## 1. Introduction

Pesticides are utilized worldwide in the agricultural and other industries in two million tons per year. In contrast to non-chemical-based pest management methods, economic and agricultural policies in many African countries encourage the use of pesticides. It will be extremely difficult for humankind to meet future food demands without further compromising the integrity of the Earth’s environmental systems in the next decades [1–7]. Although agricultural systems presently play a significant role in the destruction of the environment, population expansion and rising consumption are predicted to roughly treble that demand by 2050 [8–14]. In response to these demands, “sustainable intensification” is receiving more attention as a way to boost yields on underutilized landscapes while at the same time lowering the environmental impacts of agricultural systems [15–20]. The persistence of pesticides in the environment as a result of their accidental or deliberate usage has had an impact on ecosystems and organisms that are not intended targets. Acute and chronic pesticide poisoning typically happens as a result of eating contaminated food, having a chemical mishap at work, or being exposed to pesticides while working in agriculture [21–30]. Nigeria imports 135 pesticide compounds totaling roughly 15,000 metric tons of pesticides per year [31]. The Environmental Protection Agency (EPA) estimates that agricultural output accounts for 76% of all pesticide use in the country, with the remaining 24% going to the urban, industrial, forestry, and public sectors [32]. With less work, agricultural production has risen owing to these chemicals. However, issues related to incorrect pesticide use had resulted in human disease, wildlife extinctions, and deterioration of water quality [21-30, 32]. Research on pesticides are seen to be crucial for lowering pesticide risk and enhancing public health regulations [33–38]. In Nigeria, it has been calculated that between 125,000 and 130,000 metric tons of insecticides are used annually [39]. Nonetheless, the use of pesticides has also given rise to worries about their impact on the environment and the possibility of harmful or cancer-causing residues persisting in the food chain [1-3]. Investigations revealed that the usage of some deadly chemicals during the production of the yam flour may be blame for cases of food poisoning that affected three households in Kano [40]. Another case of food poisoning in five families in Ilorin, central Nigeria, was linked to the intake of yam flour [41]. The usage and abuse of agrochemicals and pesticides on cereals and other agricultural goods in Nigeria has been connected to the rise in incidents of food poisoning. According to the research, these pesticides were misused and used incorrectly in order to avoid pests. Twenty thousand fatalities and over three million episodes of acute food poisoning are caused by exposure to food pesticides each year. A sudden development of gastrointestinal problems among attendees and diners at a funeral service led to 60 cases of food poisoning and three fatalities [42]. Food is still necessary since it contains nutrients that support life even at the cellular level. Regardless of social status, origin, gender, or age, everyone in human society is affected by the critical socioeconomic and scientific issue of food security [21–30, 43]. According to the definition of food security given by the UN Committee on World Food Security, food security is achieved when everyone has access to enough, safe, nutrient-rich, and preferred choices of all food at all times, enabling them to meet their dietary needs and lead active, healthy lives [44, 45]. This is typically not the case, however, as food is typically seen from the perspective of sufficiency, with less emphasis being placed on its nutritional content and safety [30, 46]. Grains constitute a significant component of the world’s staple diet and include pulses like cowpea and soybean as well as cereal crops like rice, maize, and millet [47]. In Nigeria, millet and maize are significant crops that are consumed in a variety of ways. A variety of regional foods, including akamu and tuwo, are derived from maize, as well as complementary baby food [48, 49]. Millet provides a significant source of calories and important elements in a similar way [50]. The majority of the time, it is prepared as a whole grain or transformed into auxiliary dishes like tuwo and fura [51]. It is advised for people with diabetes and celiac disease due to the absence of gluten and the slow-releasing carbs it contains [52]. Cowpeas, on the other hand, can be dehulled and turned into a paste that is cooked as moi-moi or fried to produce akara. Cowpeas are typically prepared and eaten straight away. In Nigeria, particularly in urban areas, rice is gradually becoming a staple food [53].

Our food systems, which include how we grow, process, and store food, are responsible for the nutritional value, quality, and safety of the food we eat [11, 54]. The need for food is expanding along with the growth of the global population. Up to 40% of food grain losses in sub-Saharan Africa occur in storage as a result of pests [55]. Food merchants should use insecticides to combat pests as their first line of defense to reduce losses [56]. Typically, this is done to limit harm and protect company revenues. Pesticides are becoming a necessary component of food storage as a result [57]. Even though pesticides are “essential” and have long been crucial to food security, their sustainable usage has generated controversy [58]. 584 tonnes of the approximately 147,446 tonnes of pesticides imported into Nigeria for agricultural usage in 2018 were reportedly dangerous [59]. Pesticides are not adequately regulated, and their presence in food has raised concerns across the globe [60]. Any organism that tampers with stored goods is referred to as a pest in the context of food storage [61]. They consist of rodents, bacteria, and insects. Insecticides, antimicrobials (fungicides and antibacterial), and rodenticides are thus some of the pesticides used on stored goods [62–64]. Pesticides can come from either natural or artificial sources. For instance, whereas botanicals or pesticides produced from plants are typically benign, synthetic pesticides used to eradicate insect pests have been mostly to blame for the presence of toxins in food [65]. Synthetic pesticides can be divided into four categories based on their chemical structures: organochlorine (OCPs), organophosphate (OPPs), carbamates, and pyrethroids [66]. In contrast to OCPs and OPPs, which have severe toxicity, pyrethroids can also be synthesized from the flowers of plants in the genus Chrysanthemum [67–69]. OCPs are classified as persistent organic compounds because they are stable despite the rapid degradation of OPPs and carbamates [70]. For instance, Dichlorodiphenyl-Trichloroethane (DDT), a form of OCP, may take up to 30 years to degrade [71]. Therefore, research has indicated that pesticides constitute a significant global cause of death. Pesticide residues in food have been associated with a number of illnesses, including cancer, allergies, irritability, self-poisoning, and problems with reproduction and birth [72]. In Nigeria, there have been reports of low compliance with pesticide restrictions, ignorance of the risks associated with pesticide exposure, inadequately enforced laws, and a lack of efficient monitoring programs for locally produced and consumed food [49, 73]. In order to identify and track potential health risks linked with pesticide usage and its effects on health, it is crucial to research pesticide residues in food. Additionally, it would offer crucial data or proof that decision-makers in governmental and non-governmental groups would need to take action. This study’s goal is to evaluate consumer awareness of pesticide use and common pesticide residues in grain-based foods (brown and white beans, yellow and white maize, brown millet, and rice) in a few Southwest Nigerian markets.

## 2. Material and Methods

### Study Area

The study area is selected markets across four states (Ekiti, Oyo, Osun and Ondo States) in Southwestern, Nigeria. The markets were: Oja-titun (market) Ile-Ife (16° 18′ N 23° 33′ E), Osun; Alesinloye Market Ibadan, Oyo (7° 26′ N 3° 55′ E; Oja Oba, Ado-Ekiti, Ekiti (7° 37′ N 5° 13′ E); and Oja Oba, Akure Ondo State (7° 15′ N 5° 12′ E) (see Fig. 1 below). Hereinafter, the market will be referred to as Ekiti, Oyo, Osun and Ondo, for clarity.

**Fig. 1:**
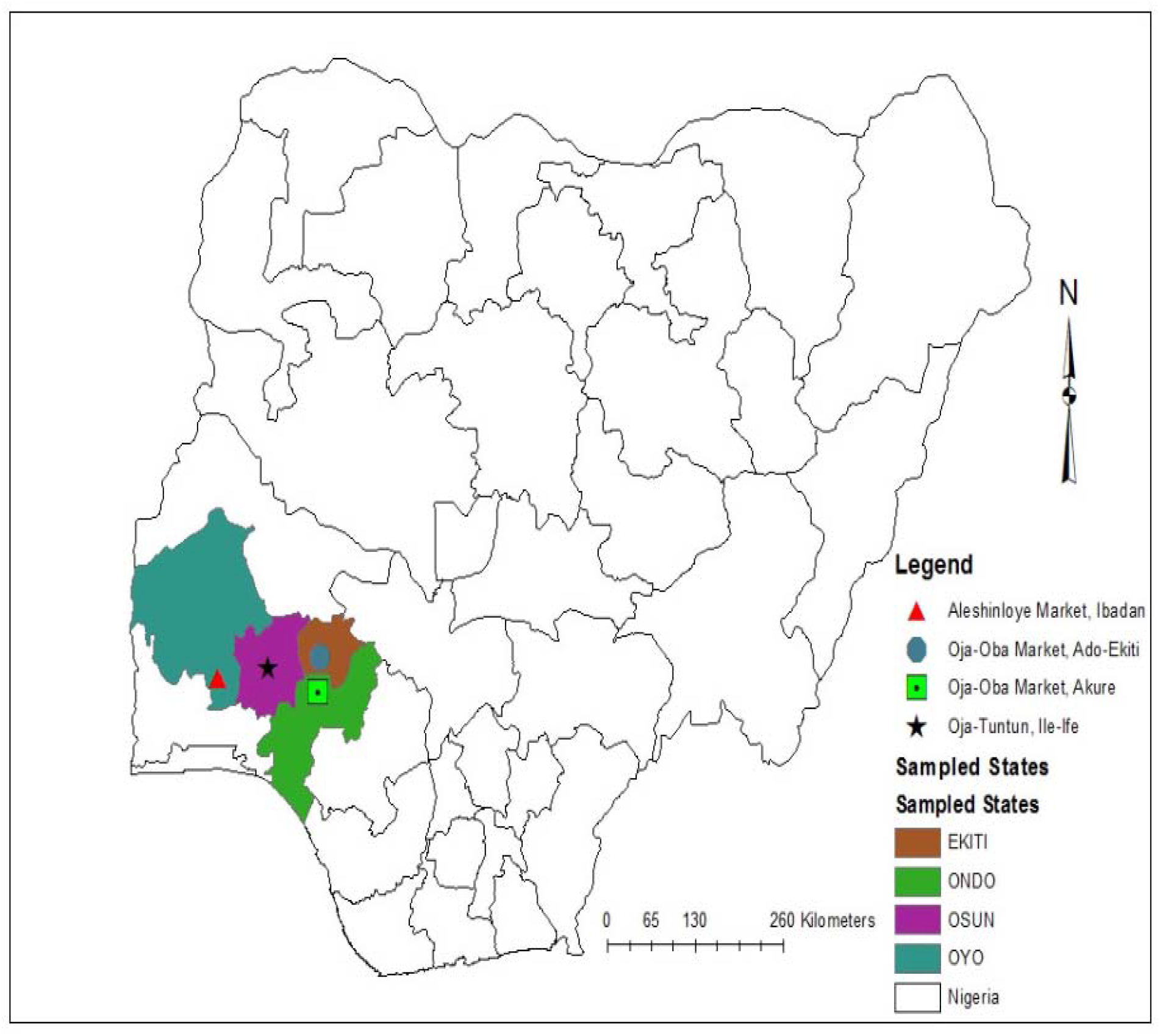
Map showing the study area

### Sample collection and preparation

Samples of dry beans, maize, millet, and rice (local and foreign) were purchased from the four markets. Six samples of brown and white beans, white and yellow maize, brown millet, and rice were collected from all four markets except for Ondo (Oja-Oba market, Akure) where yellow maize was not available. The grain samples were bought in the dried state such that there was no need for the samples to undergo additional drying. We patronized grain merchant and purchased each of the six grains sample. Each grain sample was bought in three separate portions of 200 – 250g and put in black-coloured polyethylene plastic bags, labelled and transported to the laboratory. On arrival, the samples were sorted to remove impurities including stones and shafts. Thereafter, the samples were thoroughly ground using mortar and pestle, and thereafter a hand-grinding machine was used to pulverize into fine powder. Finally, each of the powdered grain sample was stored in labelled Ziploc bag and kept at 4°C in a refrigerator.

### Proximate analysis

The grain samples were subjected to proximate analysis including ash content, moisture content, crude fat, crude fibre, crude protein, and total carbohydrate [74]. Triplicate samples were constituted by randomly picked samples from three of the four markets. The replicates of each market were pooled.

### Determination of moisture content

Moisture content was determined after drying using the oven. First, the crucibles were cleaned thoroughly and dried in an oven. After cooling, the weights of the crucible were recorded. Thereafter, 1.0g of each sample was weighed and put into the crucible before drying at 105^°^C until constant weights were attained. Moisture content was calculated using the formula:

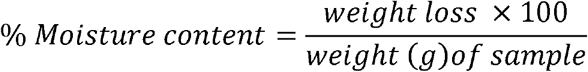

### Determination of ash content

To determine the ash content, each finely ground samples was weighed into a clean, moisture-free crucible. The organic matter was charred by burning over low flame without the lid covering. Thereafter, crucible was transferred to a muffle furnace at 600^°^C for 6h. Following this, the samples became completely ashed. It was then cooled in a desiccator and reweighed.

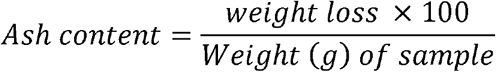

### Determination of crude protein

The micro-Kjeldah method was used in determining the crude protein of the samples [74].

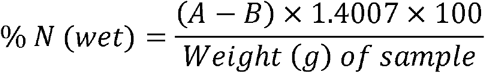

A is the volume (ml) of std HCl × the normality of std HCl

B is the volume (ml) of std NaOH × normality of std NaOH

### Determination of crude fat

The crude fat content of each sample was determined using Soxhlet apparatus. 10g of each sample added to a pre-weighed filter paper. The filter paper was folded carefully, moved into a pre-weighed thimble and reweighed before the transfer into the Soxhlet apparatus for extraction using n-hexane at 60 – 80ºC for 6 hours. Thereafter, the solvent evaporated after exposing to 100ºC in an oven for half an hour. On cooling (in a desiccator), the thimble was reweighed. The fat extracted from a given quantity of sample was then calculated. This was carried out gravimetrically as 5g of the sample was weighed into thimble. Petroleum ether was used for extraction at 60 – 80°C for 3 hours. The solvent was evaporated, and the flask reweighed.

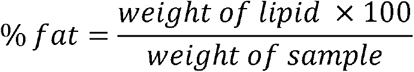

### Determination of Crude fibre

A portion of the ether extract (fatless) was used in crude fibre determination. First, dilute acid was used in the heating the ether extract This was then serially heated with dilute acid and then hydrolyzed with dilute alkali so the digestible portion is negated. Finally, the difference between the weight of the residue and the ash was recorded as the fibre content [74].

### Determination of total carbohydrate

The percentage carbohydrate content of the samples was determined indirectly from other proximate composition as follows:

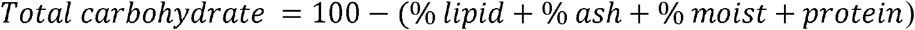

## 3. Determination of pesticide residues

### Chemicals and reagents

Commercial standards of Quinalphos, Diazinon, Fenitrothion, Dimethoate, Acephate, Malathion and Chlorpyrifos were gotten from Sigma Aldrich Laborchemikalien (St Louis, MO, USA) through Top Scientific Ltd. Ado-Ekiti Ekiti state, Nigeria. They all had >99.6% purity. Other regents including sodium chloride (NaCl), acetone, methanol, anhydrous magnesium sulphate (MgSO_4_), gradient grade acetonitrile and Primary Secondary Amine (PSA) were purchased from Top Scientific Ltd. Ado-Ekiti Ekiti state, Nigeria.

### Preparation of pesticide standard solution

The stock solutions of each of the aforementioned commercial standard reagents were prepared separately in acetone (1000 mg/L). A mixed standard solution (50 mg/L actetone) containing the standard reagents was prepared from the individual stock solution and then a lower concentration of 10 mg/L. Thereafter 0.1, 0.2, 0.5, 1.0, 2.0, 3.0, and 5.0 mg/L working standard were prepared by measuring the required volume to make up each of the concentration in a 10ml flask and making it up with the stock solution of acetone.

### Extraction and clean up

QuEChERS extraction was carried out as described by the European Committee for Standardization [75]. First, 10g of the homogenized sample was weighted into a 50 mL polypropylene centrifuge tube, 10 mL of acetonitrile (MeCN) was added followed by the use of a vortex mixer to shake the centrifuge for 30s. NaCl (1 g) and Anhydrous MgSO_4_ (4g) were added followed by immediate shaking with the vortex mixer for 60 seconds to pre-empt the MgSO_4_ from becoming aggregates and then, the extract was centrifuged at 5000 rpm for 5 minutes. A 3 mL aliquot of the MeCN layer was moved into a 15 mL micro centrifuge tube holding 600 mg of and 120 mg of anhydrous MgSO_4_ and Primary Secondary Amine, respectively. The vortex was again used to thoroughly 30s before centrifuged at 4000 rpm for 5 minutes. Finally, a 0.2 μm PTFE filter was applied in filtering 1 mL supernatant before taking in a clean vial for injection.

### Instrumentation and methodology of GC-MS analysis

A Varian 3800/4000 gas chromatograph mass spectrometer was used to analyze pesticide residues. The capillary column was set at AT-1 and a length of 30 m, while the ID and film thickness were set at 0.25 mm 0.25 μm, respectively. The GC initial temperature of the GC column was 70°C and increase to 300°C after two minutes holding time. At 300°C, it was subjected to another 7 minutes holding time. Together, the total run time was 32 minutes. Nitrogen (99.9995% purity) was the carrier gas of choice. The GC-MS interface temperature was at 280 °C and carrier gas had a constant flow rate of 1.51 ml/min. Injector and detector temperatures were set at 250°C, linear velocity was the flow control mode; and a 1μl of sample was injected in the split ratio of 30:0. The range of the MS was 30 – 800 Da. The concentration of pesticide residues in each sample were obtained following the comparative peak retention times of the samples relative to the pure analytical standards; and were represented as relative area percentage. For the identification, the spectrum obtained through GC – MS compounds were compared with the database of the National Institute Standard and Technology. As a precaution to, samples were randomly and continuously injected as a batch in order to separate technical variations from the biological. More so, the prepared pooled samples were injected into the machine at regular intervals as quality control. The concentrations of the pesticide residues were given in mg/kg.

### Survey

Structured questionnaires were administered to 60 respondents in each of the four markets. The questionnaire included questions relating to the: socio-demographic information, attitudes of grain merchants towards grains preservation, knowledge of grain merchants towards pesticide application and the health implication of pesticide usage on humans.

### Statistical analyses and calculations

Data analyses were carried out on SPSS version 21.0, IBM, USA. The differences between the treatments were evaluated using Analysis of variance (ANOVA) while Duncan’s new multiple-range test was used to separate means at (*P* < 0.05). All graphs were plotted on Microsoft Excel, 2019.

## 4. Results

Figures 2 through 18 below show a descriptive summary of all the questions that were analyzed. Of the entire respondents, up to 50.4% never read instructions on pesticide application while a shared 24.8% read them always and occasionally (Fig. 9). The number of respondents who never read the instructions were particularly high in Ekiti and Ondo while in Osun up to 97% of the respondents reads them occasionally or always.

**Fig. 2:**
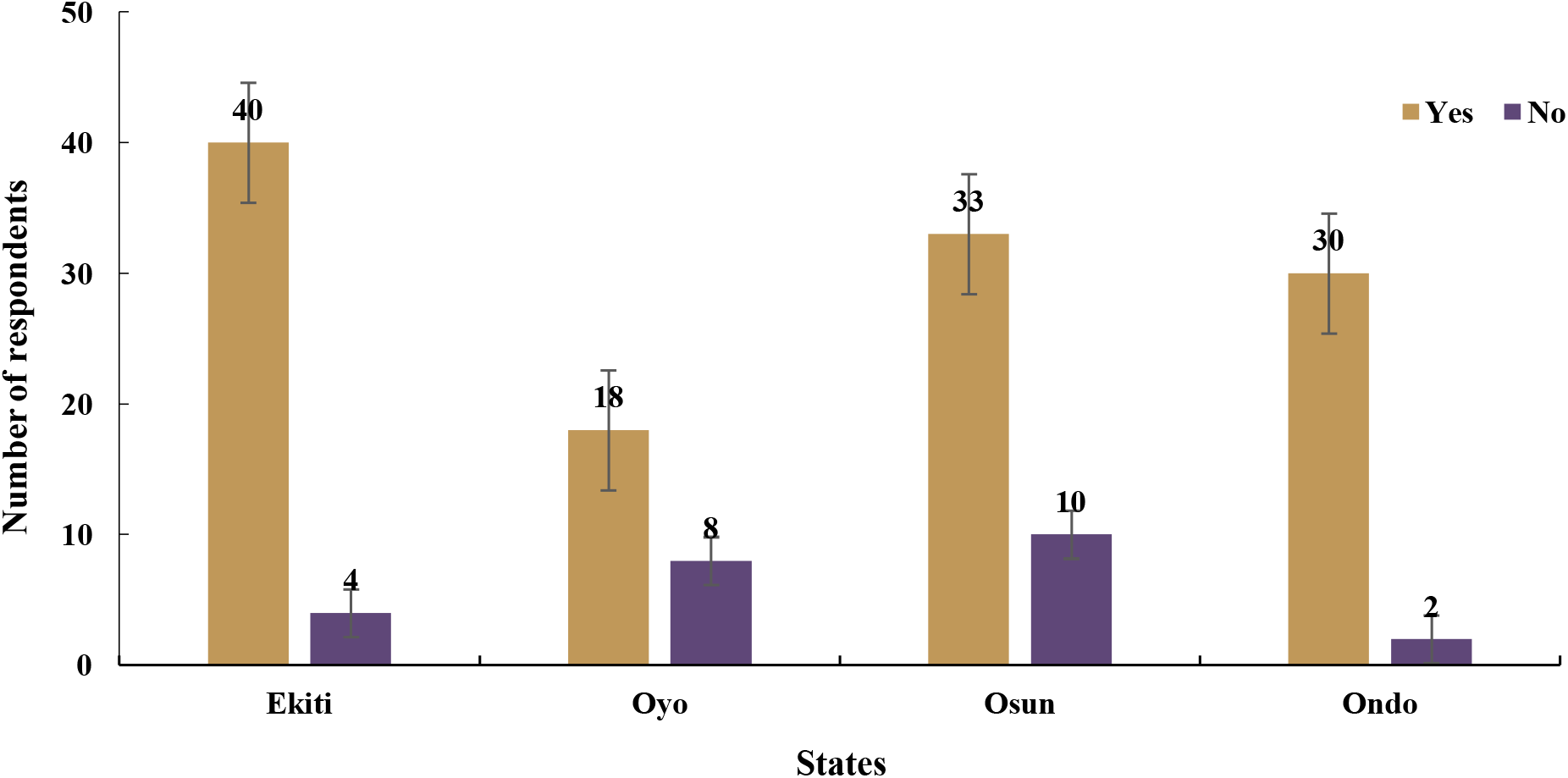
Responses of respondents to the question, do you spray your grains with pesticides to preserve it?

**Fig. 3:**
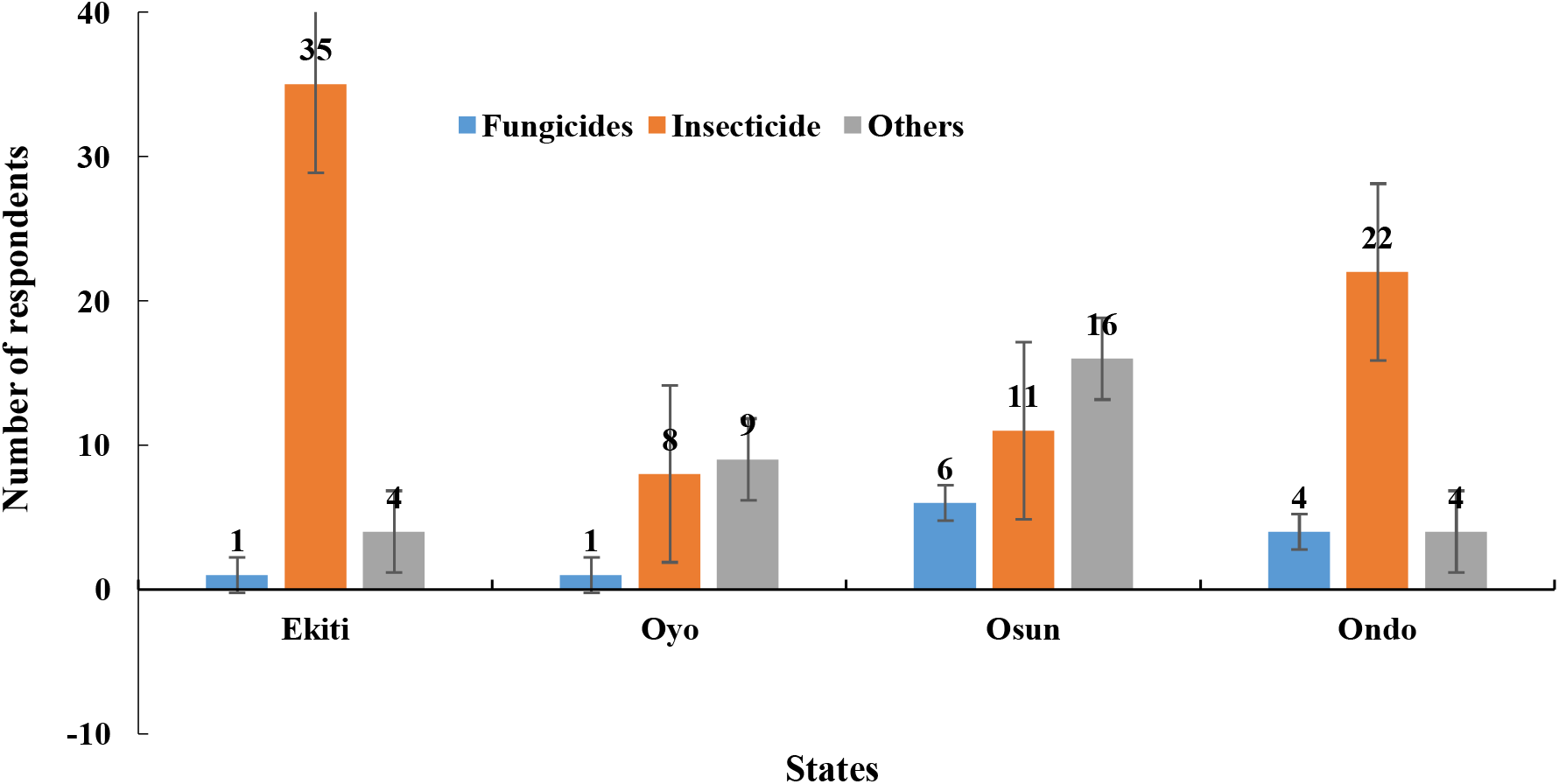
Responses of respondents to the question, which type of pesticide do you use?

**Fig. 4:**
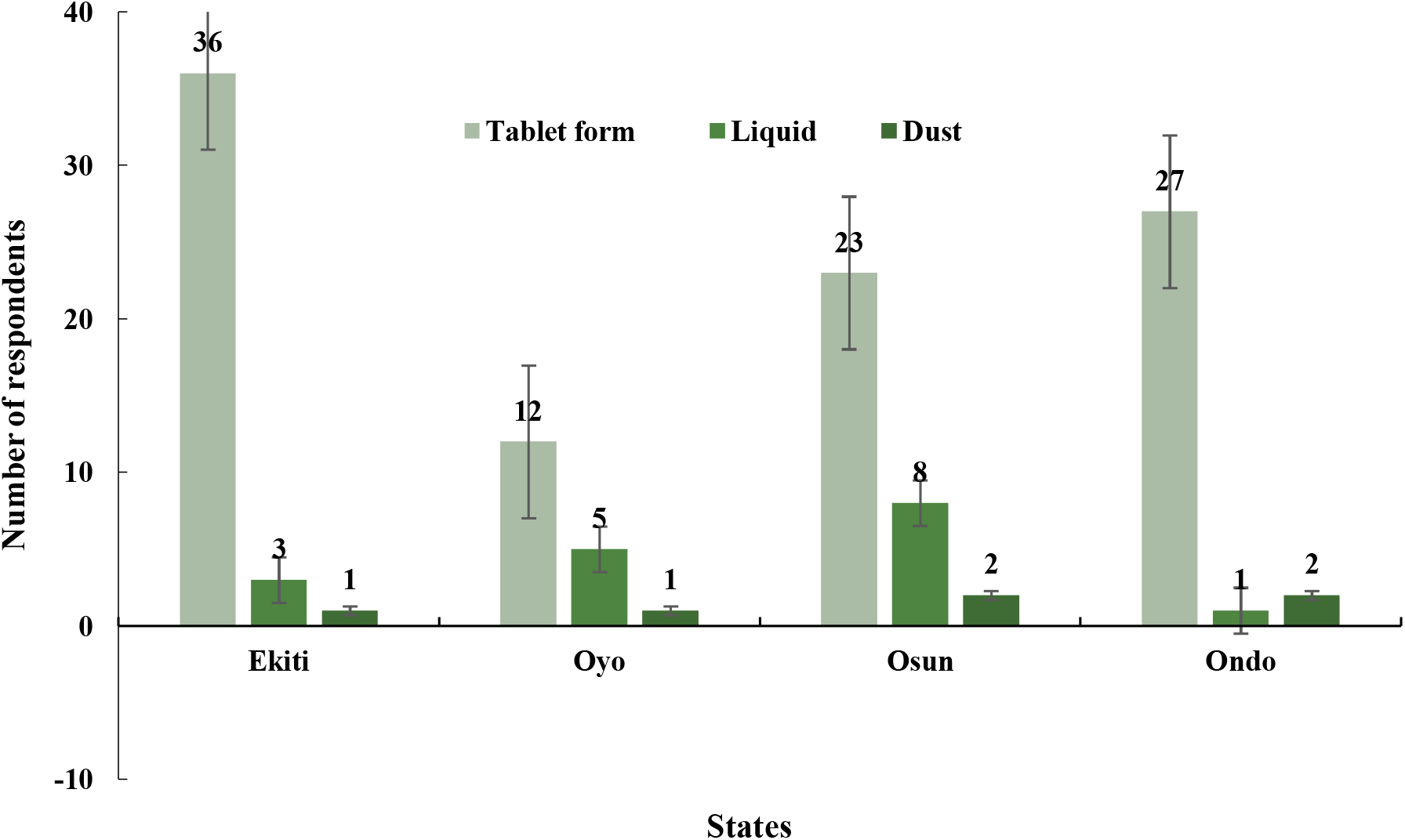
Responses of respondents to the question, which of these forms of pesticides do you use in preserving grains?

**Fig. 5:**
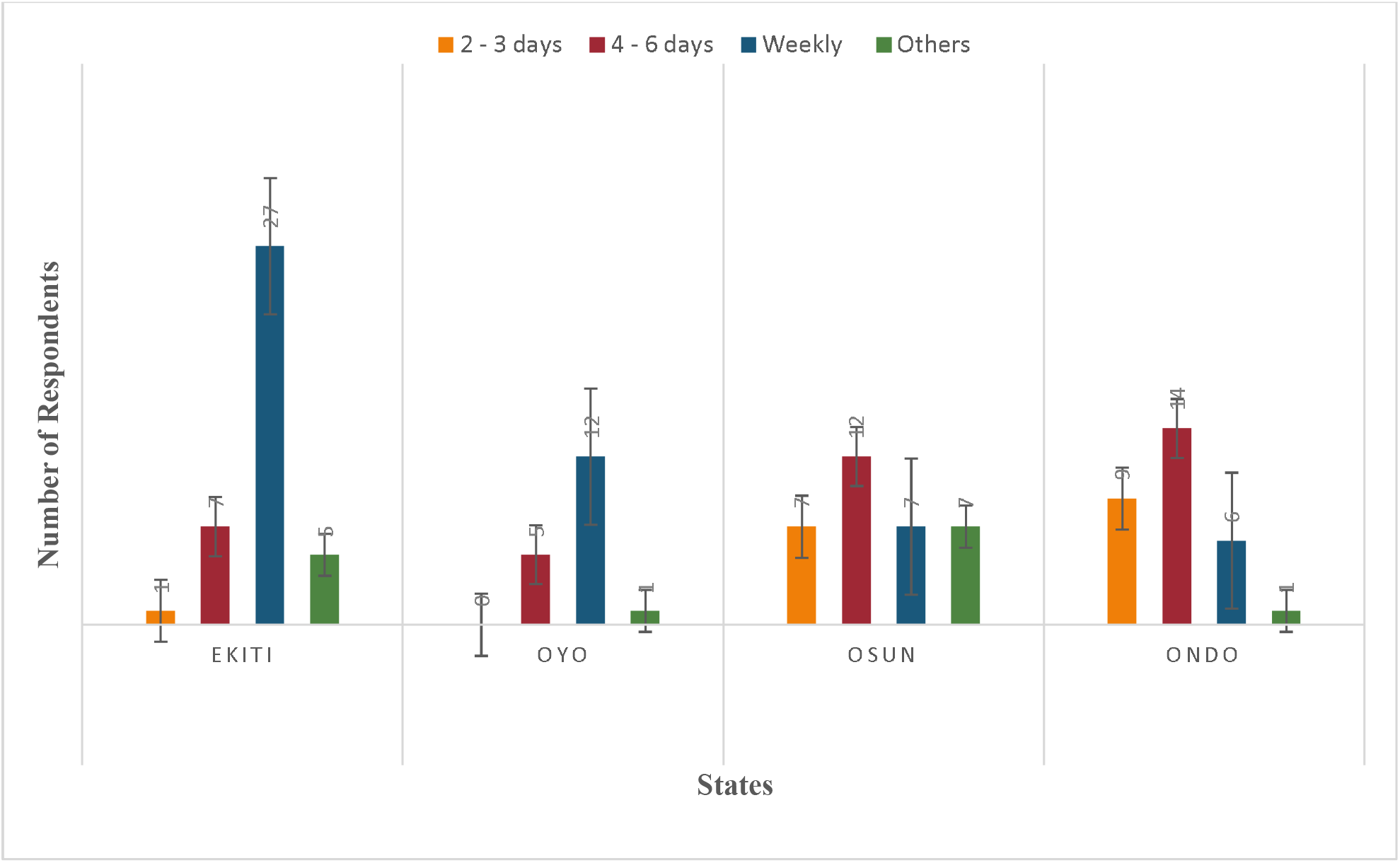
Responses of respondents to the question, at what time interval do you spray grains?

**Fig. 6:**
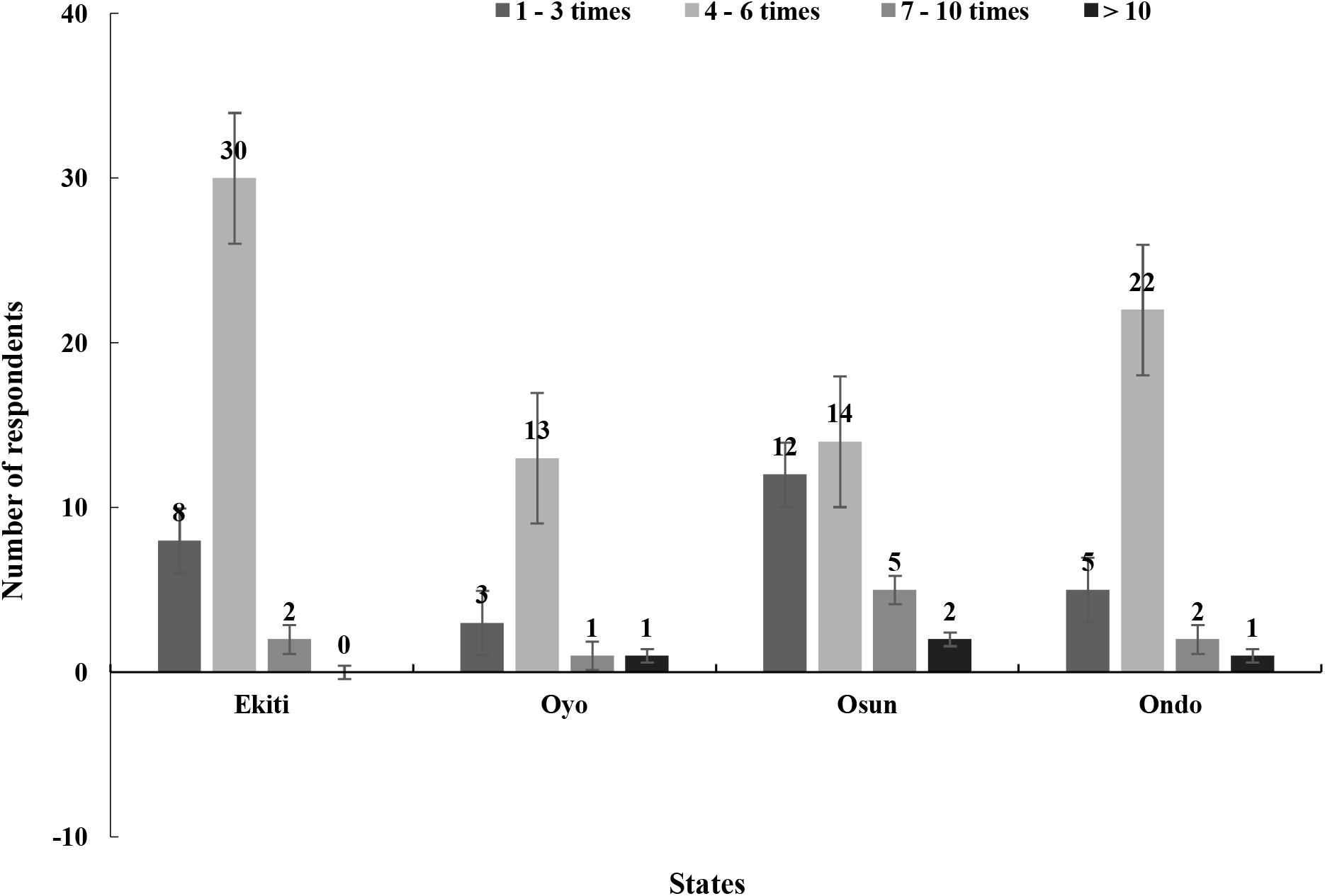
How many times do you spray grains each time?

**Fig. 7:**
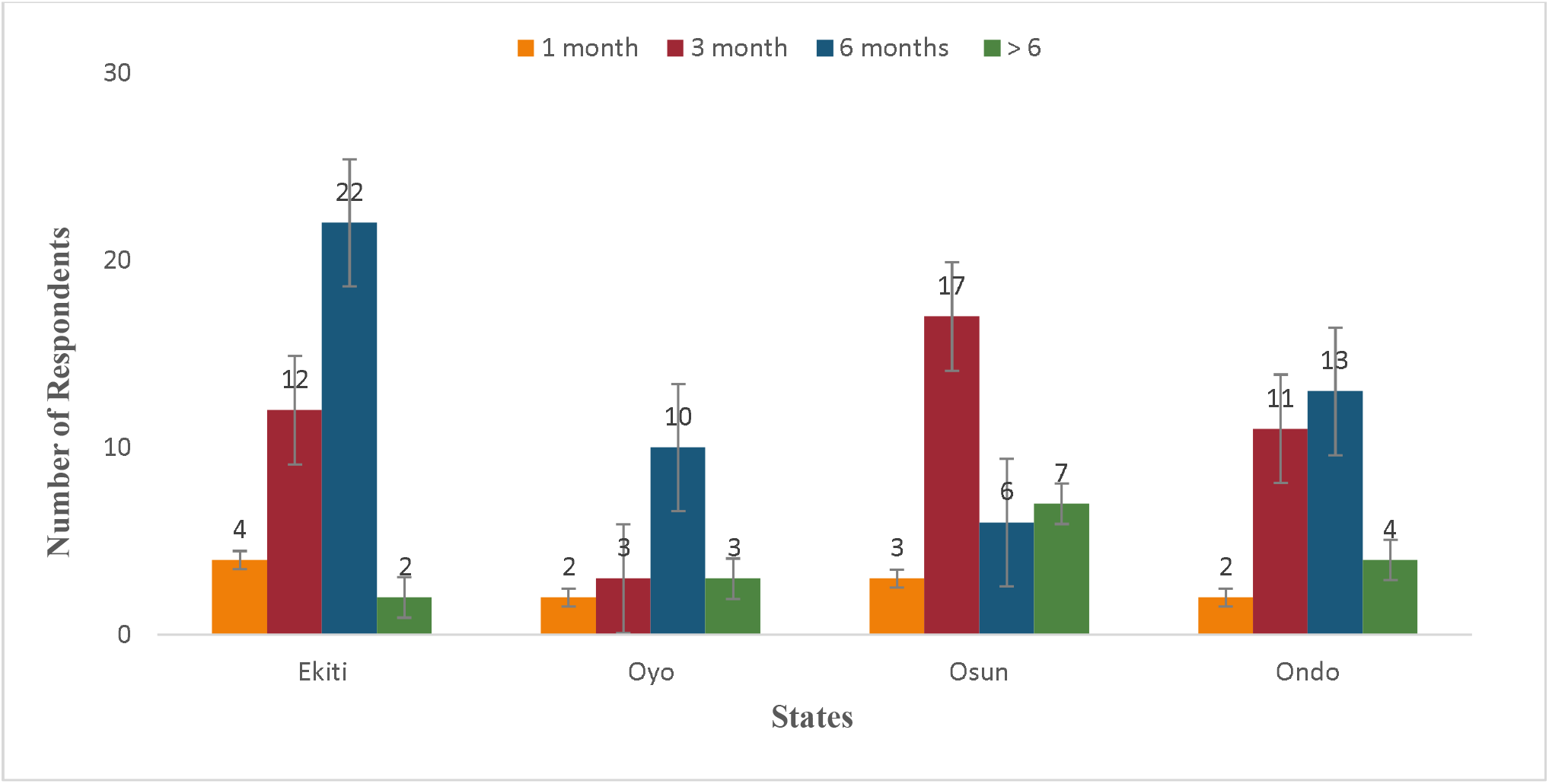
For, how long do you store your grains before selling?

**Fig. 8:**
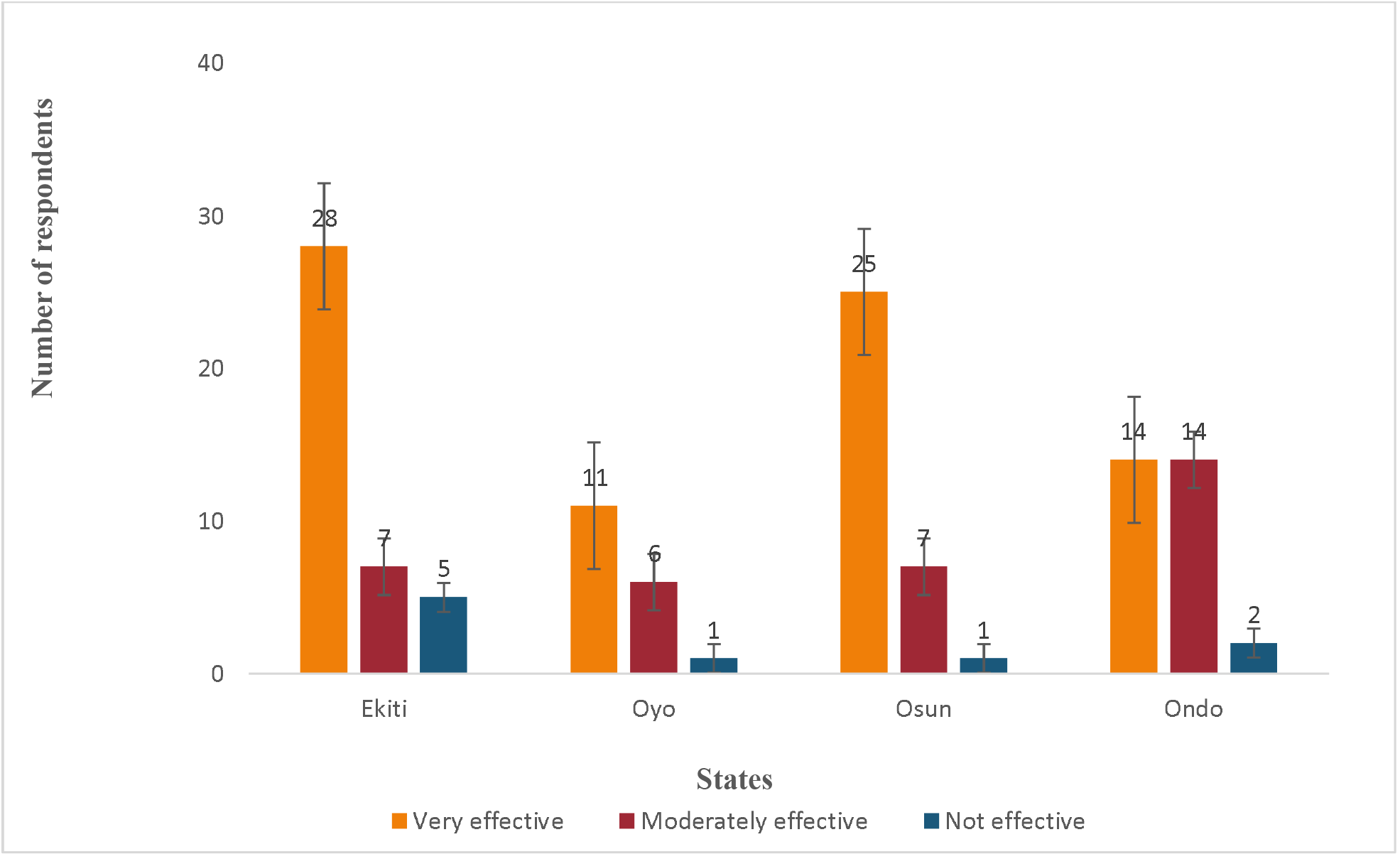
How effective are the most commonly used pesticide?

**Fig. 9:**
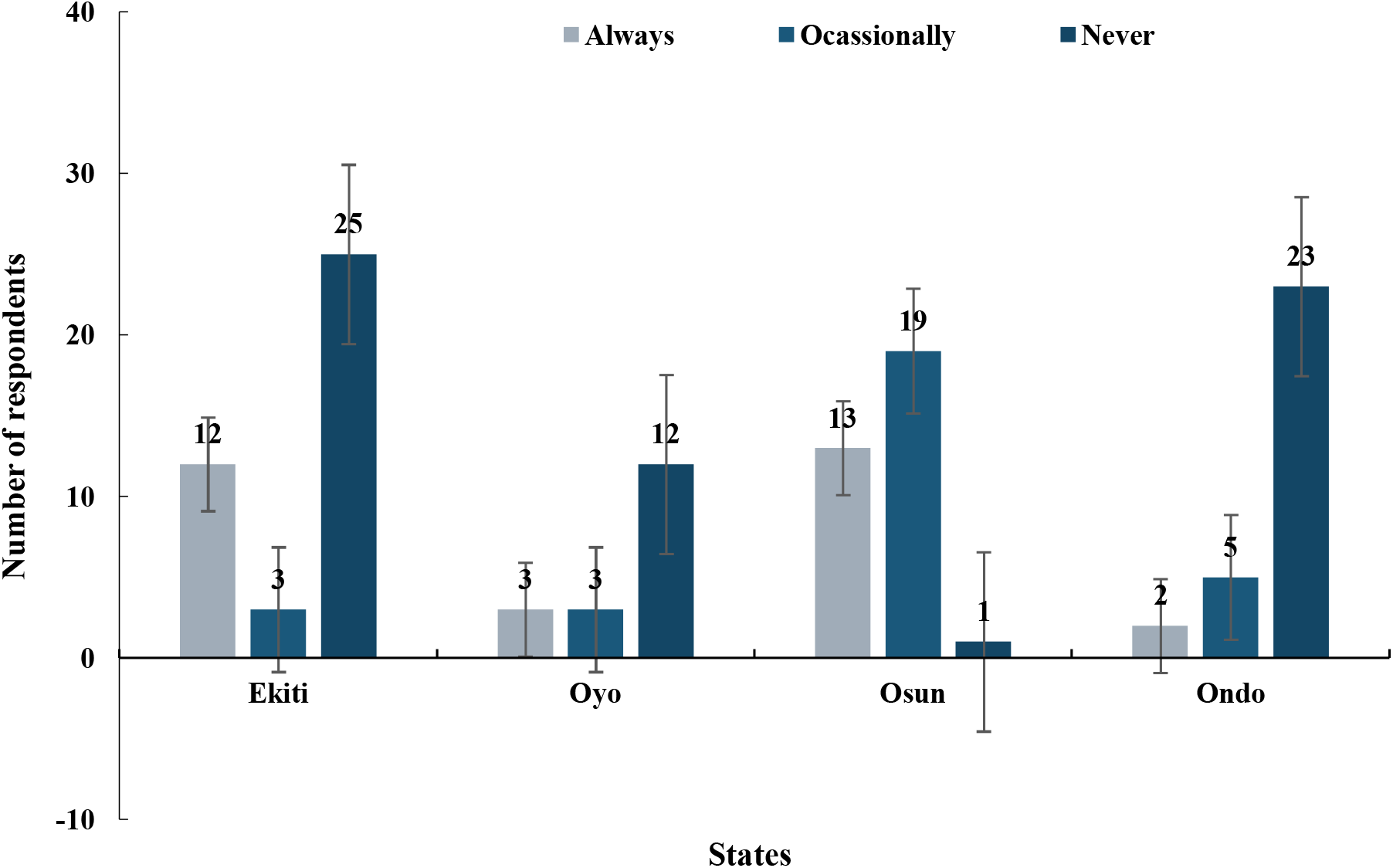
Do you read agrochemical instructions on pesticides before application?

Most respondents use PPE across all markets (Fig. 10). These were largely facemasks in Ondo, Oyo and Osun while coveralls were also widely used in Osun (Fig. 11). In Osun, Oyo and Ondo, most respondents do not get exposed to pesticides during application (Fig. 12). On the other hand, up to 52.5% of the respondents in Ekiti do not know whether or not they are exposed to pesticides during application. Across the four markets, majority of the respondents never attended any professional training on pesticide application (Fig. 13). This amounted to 79.3%, yet most of the respondents in each market think they can effectively handle pesticides (Fig. 14). Except in Ondo where 50% of the respondents were aware that unsafe pesticide application is harmful to human health, most people were not (Fig. 15). For symptoms associated with pesticide application, a wide variety of symptoms were reported by respondents following pesticide application or consumption of grains. These included Headaches and dizziness, itching and redness of the eyes, skin allergy, diarrhea and stomach disorder, vomiting and loss of appetite, weakness of the body, asthma, permanent skin patches, shortness of breath, excessive sweating (Fig. 16). Headache and dizziness were the most common symptoms and thereafter itching and redness of the eyes and weakness of the body. Asthma, diarrhea and stomach disorder were the least common.

**Fig. 10:**
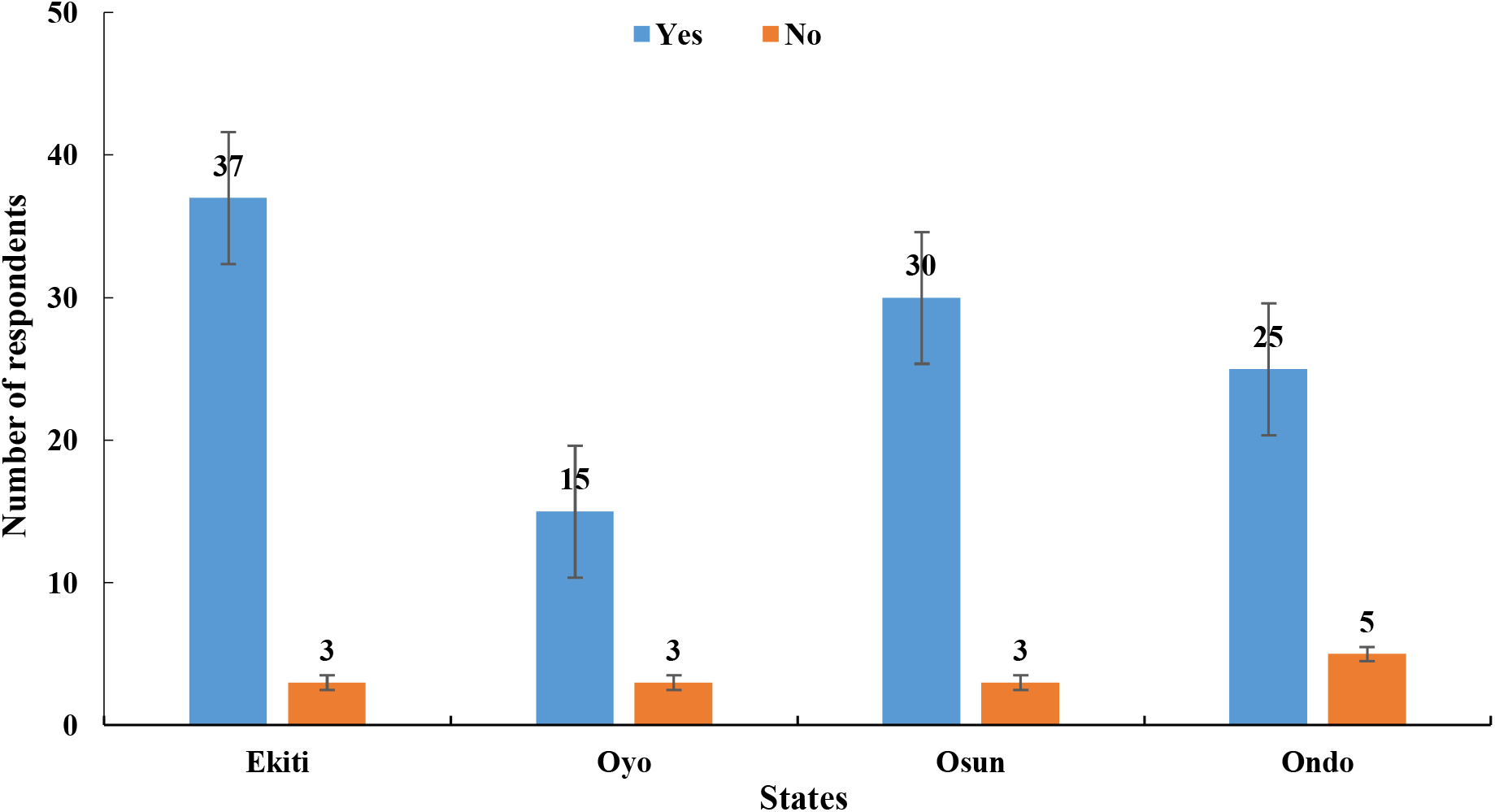
Do you use personal protective equipment when applying pesticides?

**Fig. 11:**
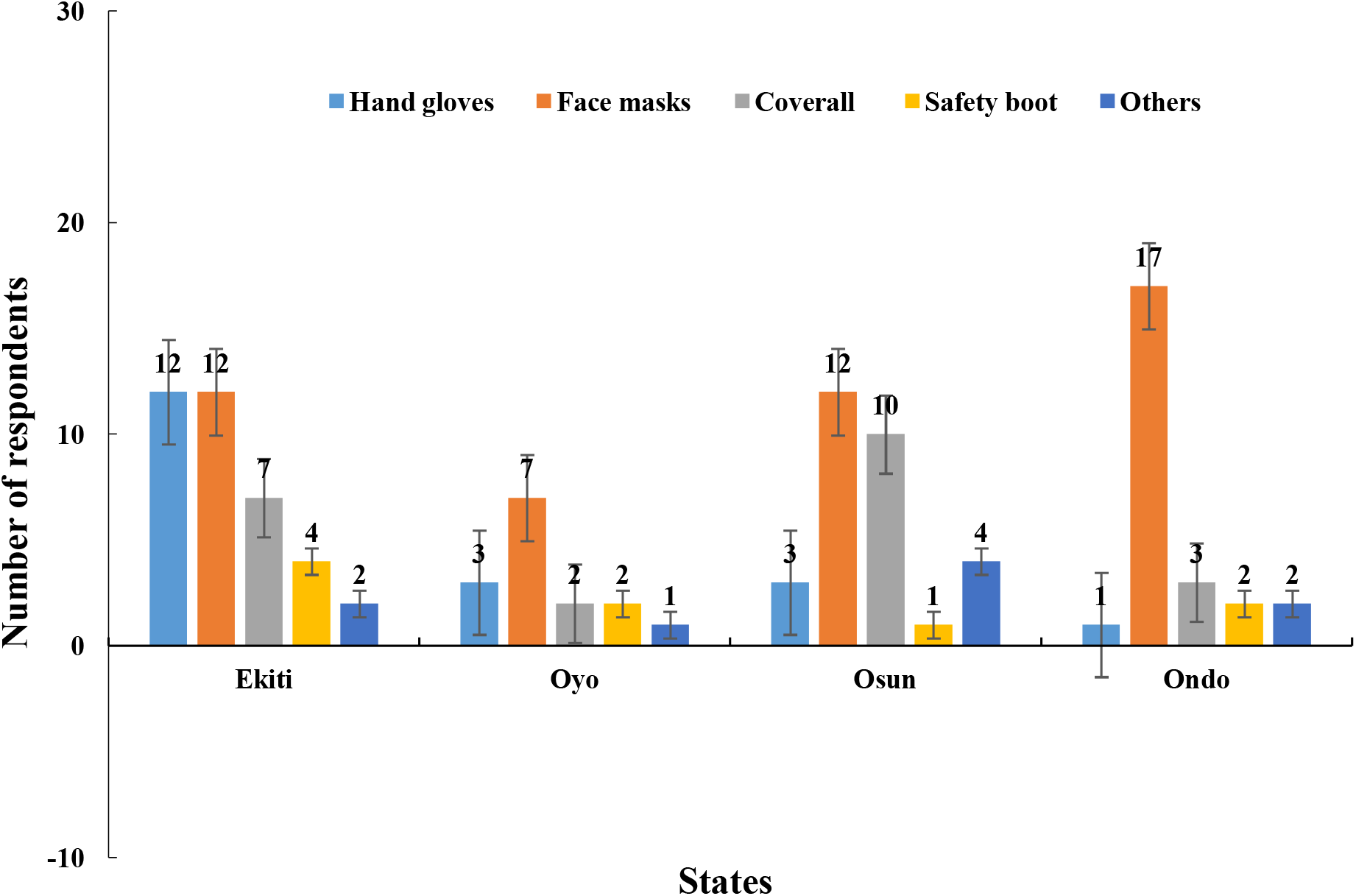
If yes, which personal protective equipment?

**Fig. 12:**
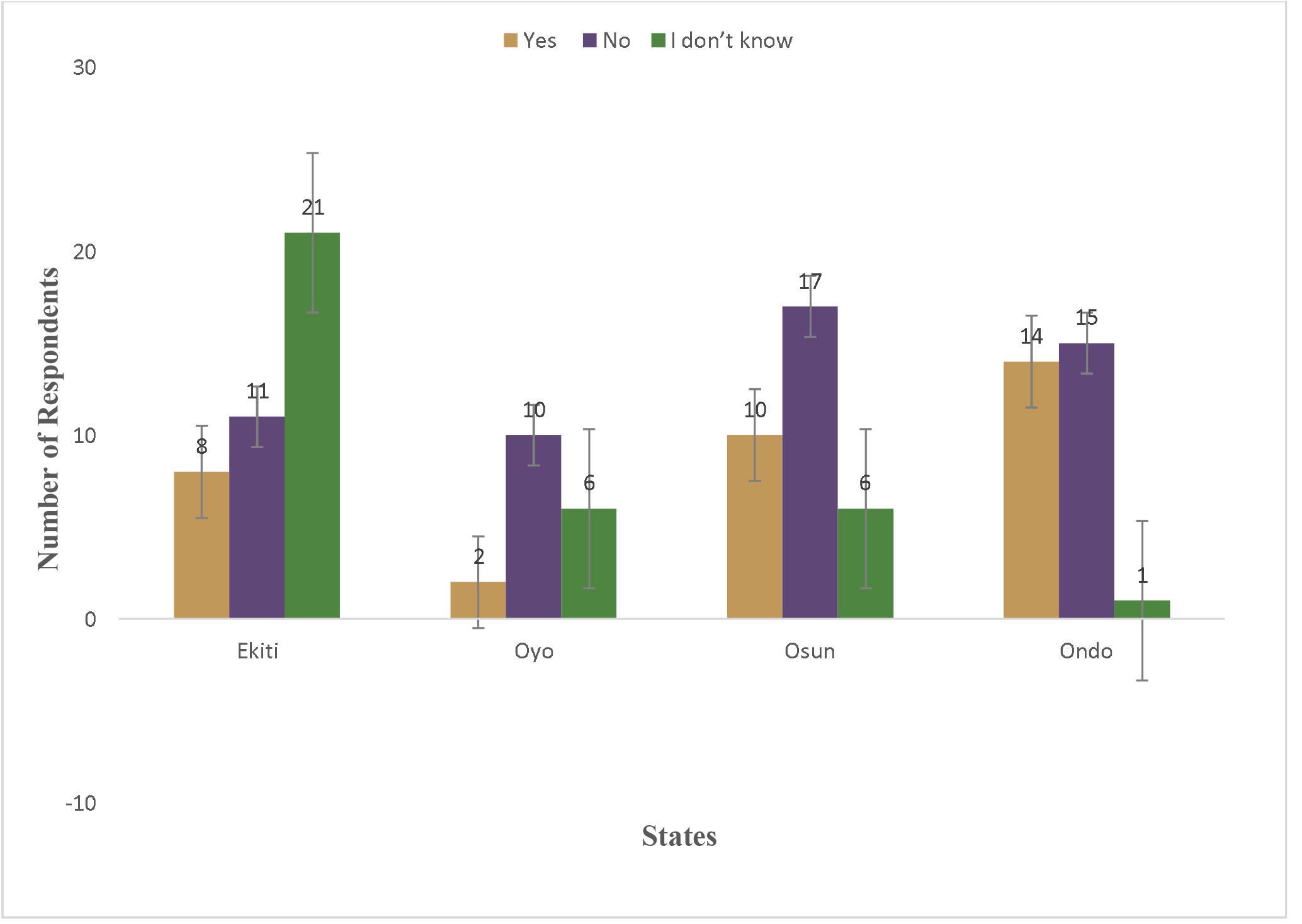
Do you accidently get exposed to pesticide during application?

**Fig. 13:**
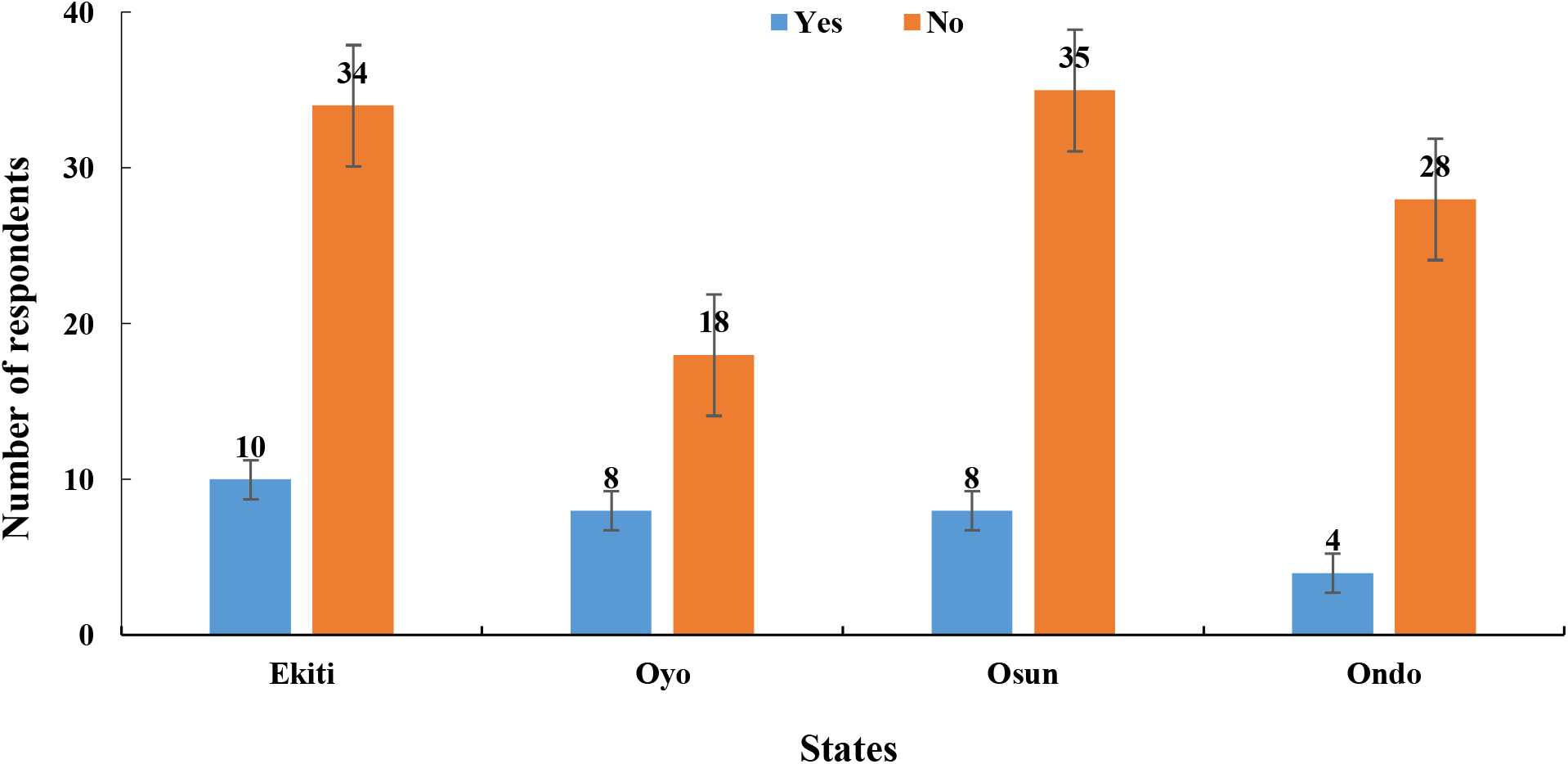
Do you attend any Professional training on pesticide application?

**Fig 14:**
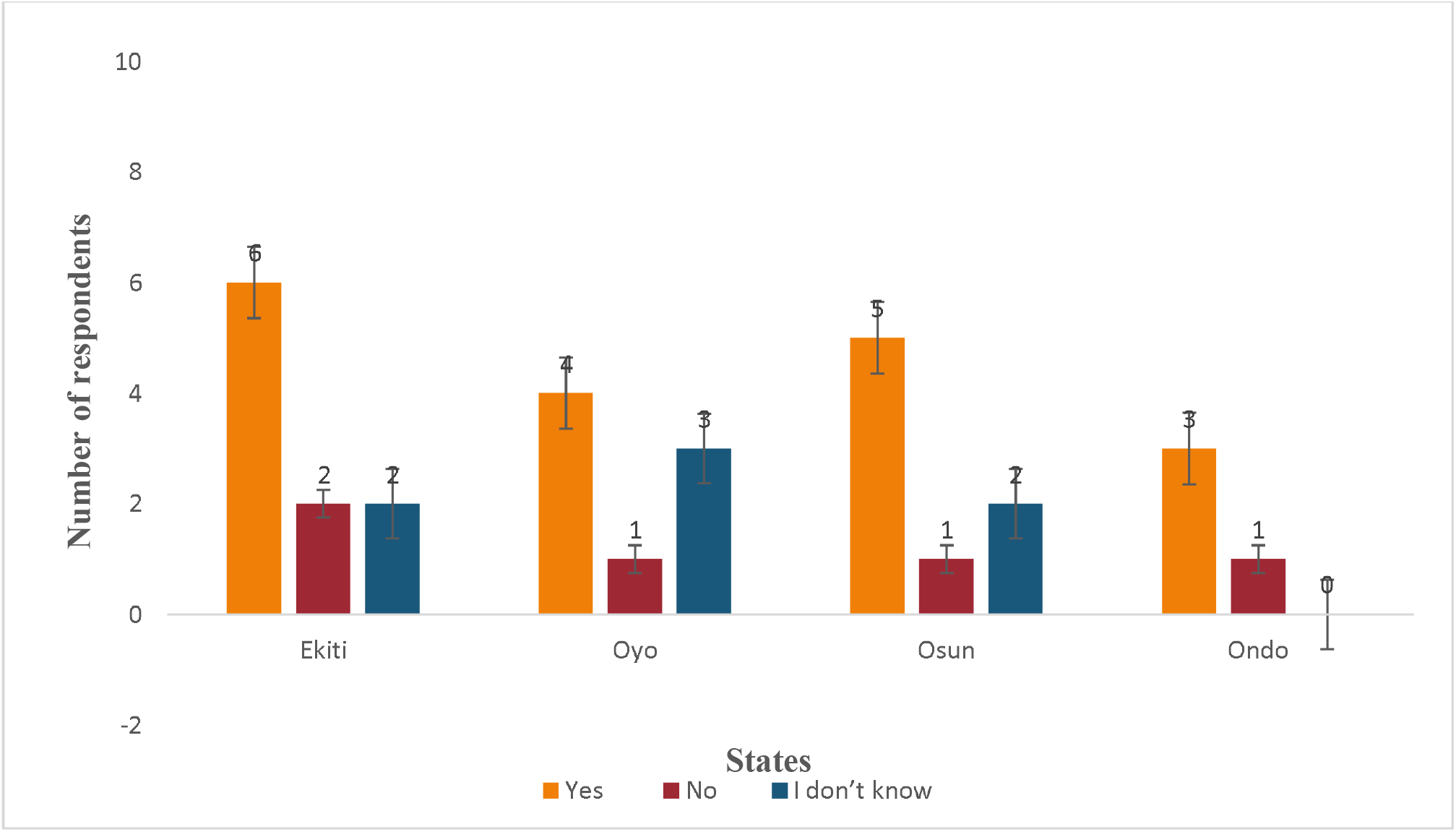
If yes, can you effectively handle pesticides?

**Fig. 15:**
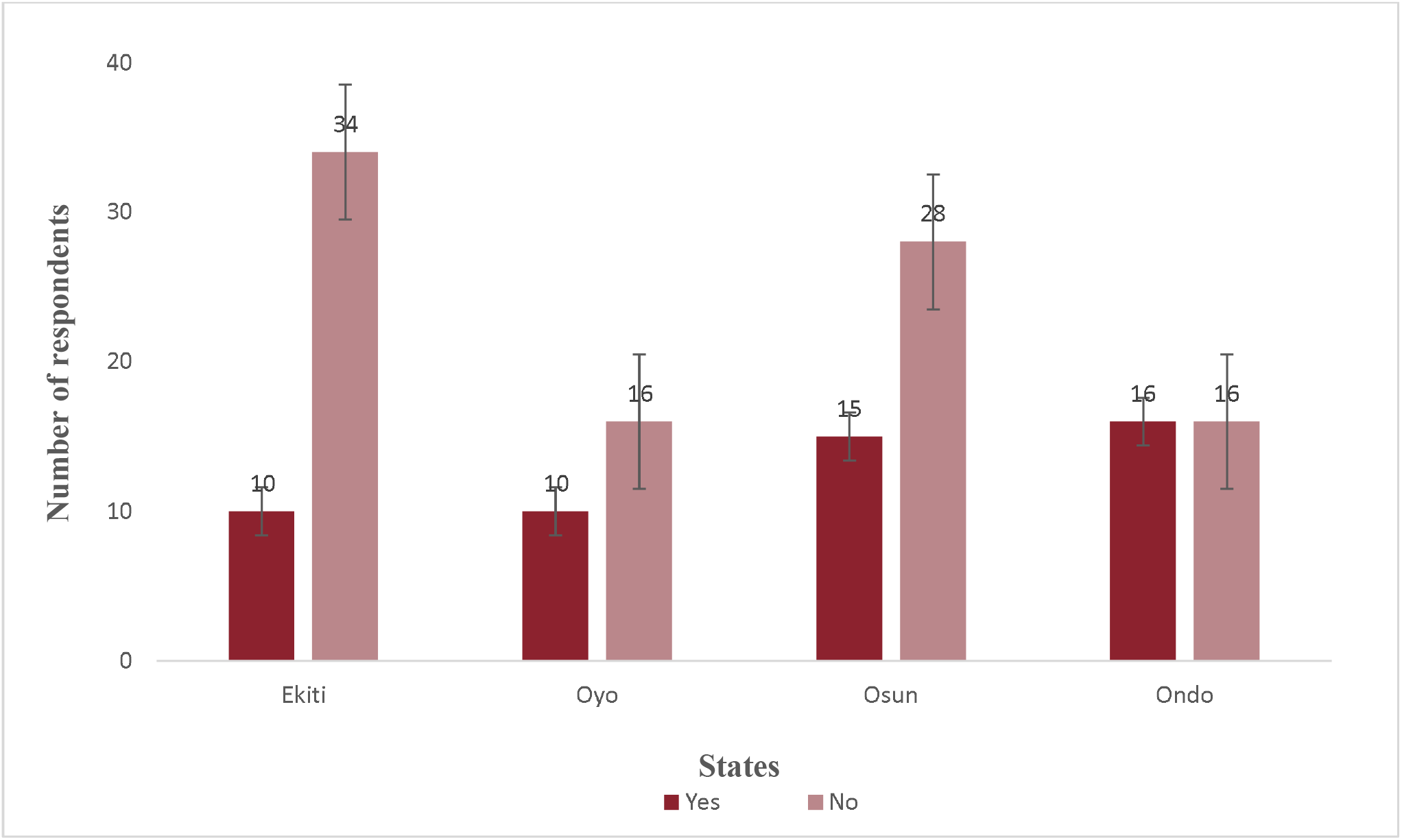
Are you aware that pesticide application of unsafe pesticide concentration is harmful to health?

**Fig. 16:**
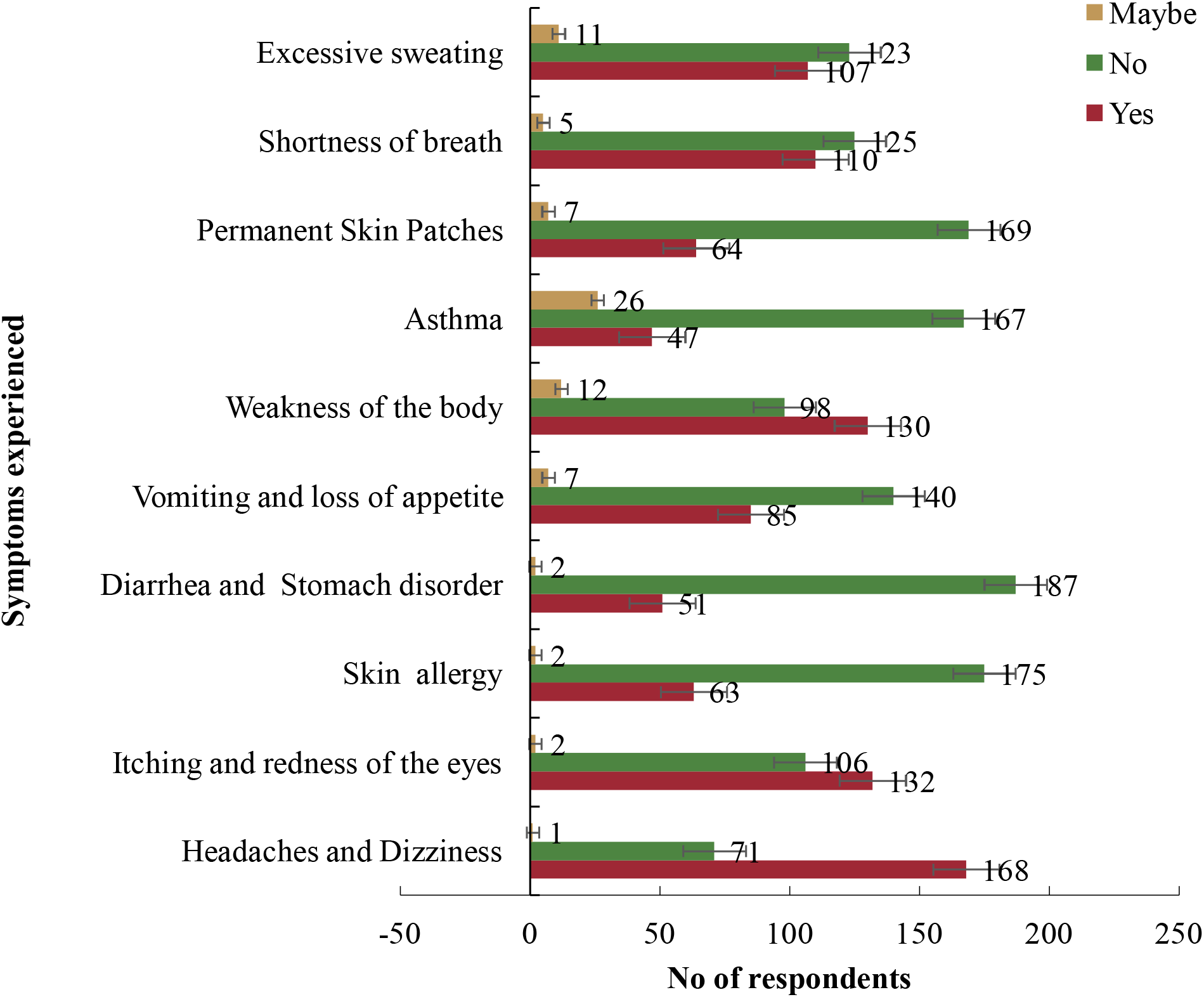
Which of these Health risks do you experience after pesticide application of unsafe pesticide concentration or after consumed grains treated with pesticides?

For common pesticides in grain, a total of 27 pesticide residues were identified in the grain samples. These were OCPs (δ-Lindane, α-Lindane, β-Hexachlorocyclohexane, p,p’-DDT, Dieldrin, p,p’-DDE, Aldrin, Endosulfan, Heptachlor and Chlordan); OPPs (Phenthoate, Chlorthiophos, Ethion, Prothiofos, Iodofenphos, Amitraz, Flumioxazin, Malathion, Dichlorvos, Pirimiphos methyl, Diazinon, Chlorpyrifos); Pyrethroids: (Cypermethrin I, Amitraz, Flumioxazin); and Carbamates (Carbaryl, Carbofuran). The percentage frequency of each pesticide group was in the order OPPs > OCPs > Pyrethroids > Carbamates (Fig. 17). Each grain crop had a total number of 16 pesticide residues. However, there were variations in the total number of individual pesticides that made up the four groups of pesticides observed in the grains (Fig. 18). Millet recorded the highest number of OCP residues while maize had the lowest. On the contrary, maize had the highest number of OPP and carbamate residues while millet had the lowest number of OPP residues. Meanwhile, maize, rice and beans had only one carbamat pesticide residue. The highest number of pyrethroid pesticide residues was jointly shared by beans and rice grains while millet had the lowest.

**Fig. 17:**
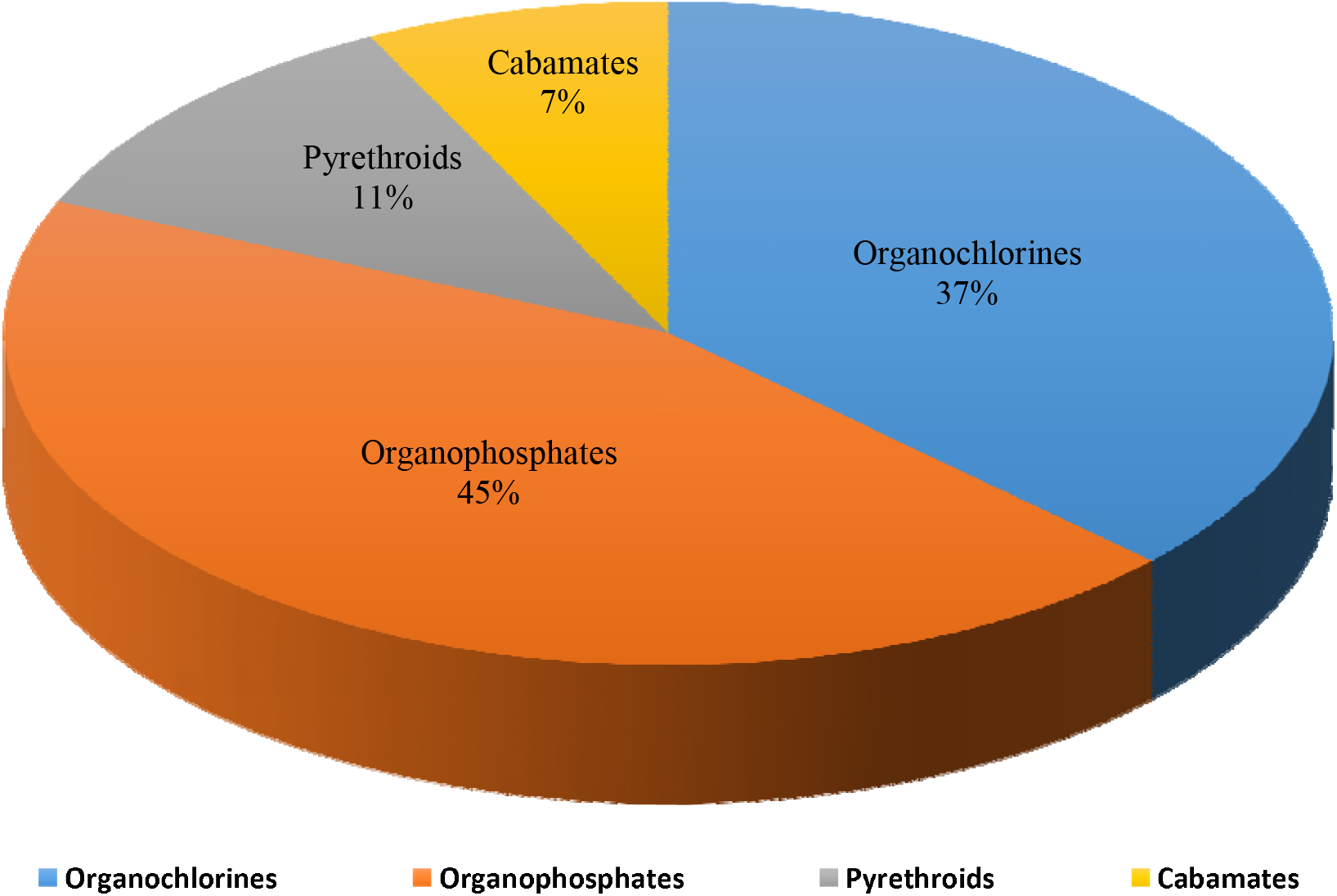
Percentage composition of total pesticide residues (27) identified in grains collected from selected markets in Southwest Nigeria

**Fig. 18:**
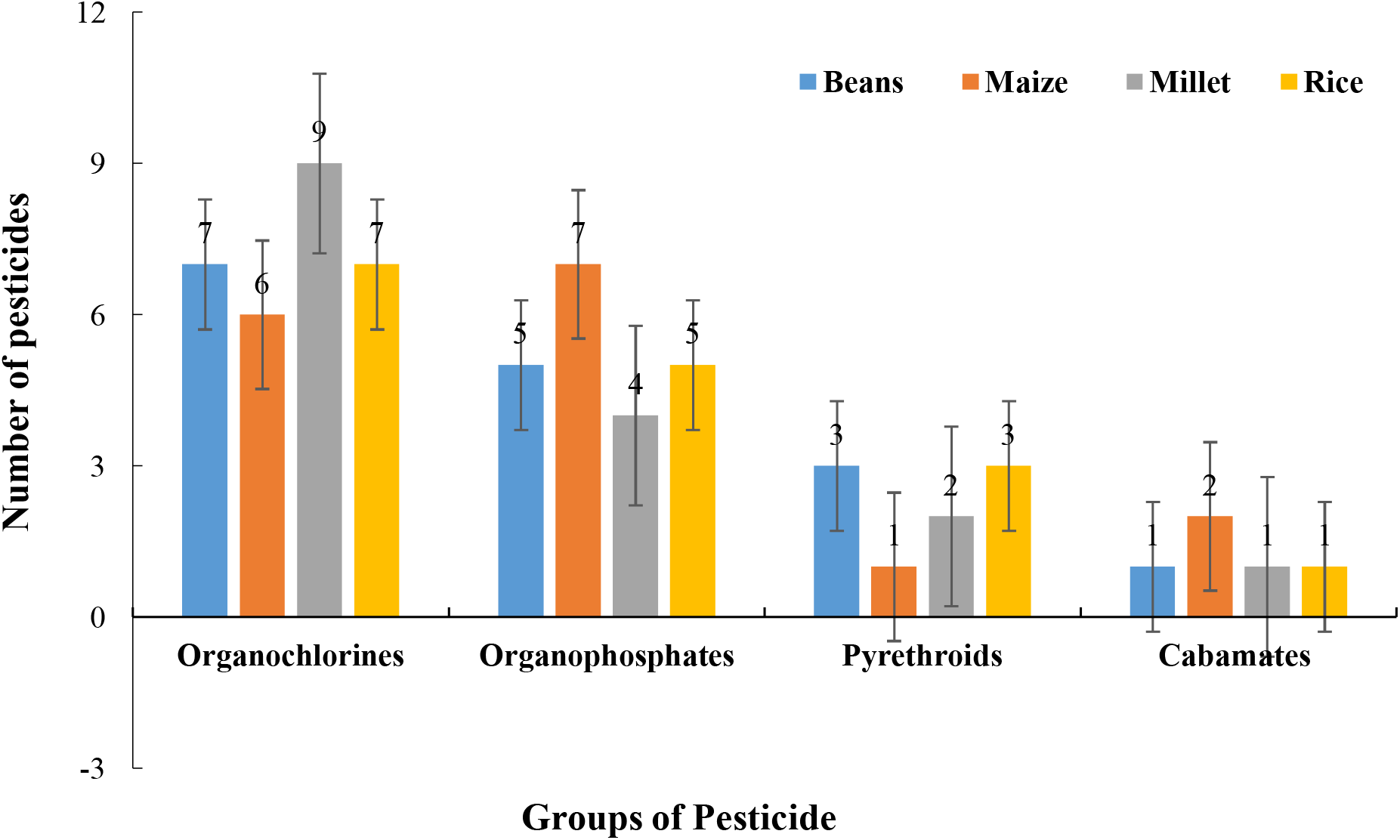
Distribution of pesticide residues in grain samples collected from selected markets in Southwest Nigeria

## 5. Discussion

### Knowledge of grain sellers on the Occurrence of pesticide residue in grains

The concentration of pesticides in grain sample is largely determined by the merchant, the type, quantity, and frequency of pesticide applied to grain during storage. The factors are directly related to the attitude and knowledge of the merchants towards grain preservation [76]. This study agrees with the study of Yusuf *et al*., [77] who state that one hundred and thirty-seven (36.5%) of the respondents that use pesticides indicated that they used phostoxin only, which is dichlorvos (an organophosphate pesticide) on their grains for storage, 88 (23.5%) indicated that they use DDForce (dichlorvos) only, 103 (27.5%) use either DDForce or phostoxin tablet (both are dichlorvos), only 11 (2.9%) use DDForce and phostoxin together, while 36 (9.6%) use “others” (i.e. Jule, Rambo, Opaayan) on maize grains. In our survey, over 80% of merchants use pesticides, mainly insecticides. This agrees with previous report that the pesticides are usually applied to food crops as the first line of action in controlling pests of stored product. The high concentration of many pesticide residues may be attributed to the many rounds of application (up to 6 times) per time. Also, the pesticides are mainly applied on a weekly basis or less across the markets. Over the storage period of 3 – 6 months observed in this study, substantial amount of pesticide residues must have accumulated. The believe that pesticides are very effective as observed in this study agrees with the findings of Alex *et al*. [78]. Overall, this suggests poor knowledge and practice of the usage of pesticides and agricultural practices by the farmers. As Mekonnen and Agonafir [79] and Tariq *et al*. [80] showed that 70% of farmers in developing countries encounter difficulties in reading instruction manuals, leading to the misuse of highly toxic pesticides. In Pakistan, the chances of pesticide misuse are relatively high for several reasons. Among farmers, there is low awareness regarding the safe use of pesticides. Furthermore, Pakistan records a low literacy rate where more than 70% of farming communities do not read or understand the national language (Urdu). To compound the problem, the awareness brochures of pesticides are printed in English and in Urdu [81, 82]. Thus, if a farmer is relatively less educated, his/her ability to absorb professional knowledge is weaker. Furthermore, his/her capability to recognize pest diseases is also weaker. Hence, the less-educated farmer tends to lack awareness of both pesticide residues and the importance of applying pesticide in standardized ways. Consequently, with less education, there is a higher chance that the farmer will apply prohibited pesticide excessively, leading to highly concentrated pesticide residues Food and Agricultural Immunology [83, 84]. In India, Agarwal and Pandey [85], Abhilash and Singh [86] found that, due to the lack of adequate knowledge regarding pesticides, farmers tend to apply the same pesticide excessively rather than mixing different kinds of pesticides rationally. In Chengdu, Li *et al*. [87] found that, due to the lack of pesticide knowledge, most farmers solely focus on the outcome of pesticide use when purchasing pesticides. They rarely pay attention to the toxic side effects of pesticides on human health. Similarly, Dongmei [88] found that the education and training provided by agricultural technology personnel affect farmers’ awareness of pesticide residues. Zhongze and Qingjiang [89] believed that a randomized security check of pesticide residues will increase the motivation of farmers to purchase low-toxicity pesticides with fewer residues or to apply pesticides in standardized ways. Furthermore, to a certain degree, the agricultural service environment and sale routes directly affect the amount of pesticides being applied, and indirectly affect farmers’ awareness of pesticide residues [90-92]. However, the intensive use of pesticides may be low on farms, but the protection of by-products, in contrast, may be high. Therefore, there is every need to train our farmers appropriately. There could also be possibilities of extraneous residues as another source of contamination because lindane and to a lesser extent endosulfan are known to be persistent in the environment and thus, capable of routinely contaminating crops and food [93, 94]. Perhaps if farmers knew better maybe food merchants would apply pesticides differently. The fact that about 50% of these merchants in our survey do not read instructions on pesticide cover explains to a large extent, the misuse of these pesticides. Although a significant proportion of merchants use PPEs, the lack of professional training and assumption by merchants that they can effectively apply pesticides makes the misuse of pesticides more likely. Similar reports of poor knowledge of pesticide application abounds [95, 96]. Our current results confirm previous works by Hussain *et al*., [26], Hussain *et al*., [27], Isah *et al*., [24]. Isah *et al*., [25], Morufu [29], Olalekan *et al*., [30], Raimi *et al*., [22] and MacFarlane’s study [97] reporting pesticide knowledge, instruction level and personal protection and Aminu and Edun [98] who reported that pesticide dose used (p<0.05), reading and adherence to instructions on pesticide labels (p<0.01).

### Common Pesticides in Grain

Agrochemicals, being toxic substances, need a correct application, because, otherwise, excessive use and incorrect selection of pesticides cause a high number of residues in the food being consumed. Residues of these substances can remain in plant tissues (e.g., fruits) and, in the long run, cause problems to human health such as cancer, depression and infertility. This control is done by the regulatory agencies of each country (e.g., NAFDAC, EPA, EFSA), and is based on the Maximum Residue Limit (MRL), varying according to the active principle and the intended culture [99]. However, the current outlook is that the world population will continue to increase, further expanding the demand for food [100]. Therefore, pesticide use in urban areas is a major concern for the aquatic environment as well as for human health and ecosystems [101]. The influence of water contamination resulting from urban pesticide runoff is greater on an aquatic environment than on human health, and food contamination is due to agricultural applications of pesticides. From this study, a total of 27 pesticide residues were identified in the grain samples. These were OCPs (δ-Lindane, α-Lindane, β-Hexachlorocyclohexane, p,p’-DDT, Dieldrin, p,p’-DDE, Aldrin, Endosulfan, Heptachlor and Chlordan); OPPs (Phenthoate, Chlorthiophos, Ethion, Prothiofos, Iodofenphos, Amitraz, Flumioxazin, Malathion, Dichlorvos, Pirimiphos methyl, Diazinon, Chlorpyrifos); Pyrethroids: (Cypermethrin I, Amitraz, Flumioxazin); and Carbamates (Carbaryl, Carbofuran) this is in tandem with the result of Alex *et al*., [78] who in his findings identified thirty (30) pesticides residues in grains, the thirty pesticide contains 90% of chemicals identified in this work. Also the study of Yusuf *et al*., [77] who state that one hundred and thirty-seven (36.5%) of the respondents that use pesticides indicated that they used phostoxin only, which is dichlorvos (an organophosphate pesticide) on their grains for storage, 88 (23.5%) indicated that they use DDForce (dichlorvos) only, 103 (27.5%) use either DDForce or phostoxin tablet (both are dichlorvos), only 11 (2.9%) use DDForce and phostoxin together, while 36 (9.6%) use “others” (i.e. jule, Rambo, opaayan) on maize grains. However, the study of Oguntade *et al*., [102] is contrary stating that grain samples collected from all the three main markets from did not contain synthetic pesticide residue as envisaged, however many bioactive substances which have pesticide activity were detected in the grain samples from various market with n-Hexadecanoic acid, Hexadecanoic acid, methyl ester and 9,12-Octadecadienoic acid (Z, Z)-, methyl ester found in all the samples.

## 6. Conclusion

Pesticides are a large and expanding part of the contemporary technology that is widely used in Africa. While worries regarding the harmful effects of pesticides on human health and the environment in developing countries have been voiced for more than ten years. The evidence provided describes a situation whose intricacy is challenging to address using the conventional methods. A crucial first step in bringing about major change in preventative health is realizing the intricacy of the processes that underlie these people’ susceptibility to pesticide exposure. The acceptance of preventive recommendations and the likelihood of their implementation will be aided by adopting a thorough viewpoint of the issue’s various elements. This study offers an intriguing update of earlier research on substantial pesticide residues. Because they are unaware of the dangers posed by pesticide residues, the recommended dosage levels, or the uniformed standards governing pesticide application, farmers frequently apply pesticides excessively and unreasonably. Problems with pesticide residue originate from this. For instance, many farmers keep upping the amount of pesticides they use despite not fully comprehending the leaking issues with sprayers. Because local health officials rarely diagnose symptoms in relation to exposure and are not trained to recognize the negative effects of pesticides, the expenses of health problems caused by pesticides, particularly medical costs, are typically not accounted for. Pesticides must be used correctly at this time to safeguard our environment and any potential health risks linked to them. These findings from the study show that farmers who only have access to information from agricultural extension officers should receive substantial and immediate attention in order to raise their level of awareness. Additionally, extensive IPM training programs must be created with the intention of disseminating precautions for protecting human health and a healthy agro-ecosystem. In order to find more effective pest management methods that utilize less pesticides, it is crucial to reevaluate the pesticide residues and common pesticides found in grains in the targeted markets. In order to lessen the harmful effects of pesticides on our environment, community development and various extension initiatives that might inform and encourage farmers to embrace the cutting-edge IPM tactics are crucial. Further research should concentrate on farmer behaviors in connection to pesticide use, an investigation of the variables that affect farmers’ awareness of pesticide residues, and regulation of pesticide availability. Research and development (R&D) should be strengthened further and the use of pesticide residue detecting technology should be encouraged in the South-West region. An alternative would be to introduce pertinent, cutting-edge foreign technologies related to pesticide residues. Pesticide residues in South-West agricultural goods can be further monitored using cutting-edge detection equipment. At the same time, it is possible to encourage farmers and the general public to form fact-based opinions about the risks posed by pesticide residues.

## 7. Recommendations

The following are hereby recommended:

I. Training in pesticide application and sensitization to the negative health effects of pesticide misuse are necessary for farmers, grain traders, and other participants in the grain value chain. This will aid in guiding their selection of pesticide, as well as the amount and timing of sprays.
II. It is important to promote, encourage, and reward the use of natural pesticides such dried lime (Citrus aurantifolia), dried neem leaves (Azadirachta indica), and dry chilli pepper (Capsicum annum). This will lessen reliance on and use of synthetic insecticides.
III. More research funding should be made available and used to investigate plants with insecticidal qualities in order to create natural insecticides. It is necessary to separate more potent chemicals from plants and turn them into simple-to-use biological or organic pesticides.
IV. Organizations that work in the food industry, like NAFDAC, need to improve their laboratory ability to monitor pesticide levels in grains, especially those that are unbranded. This will enable the tracking of dangerous grain sources.
V. The new and current restrictions on the application of pesticides should be implemented clearly and correctly. There should also be regular audits and sanctions for retailers who violate the law. Punishments for criminals will prevent others from breaking the law.
VI. The general people has to be made aware of the risks of pesticide exposure and counseled against purchasing unbranded and uncertified grains from marketplaces. This will protect them from the risks associated with pesticide use.

## Data Availability

All data produced in the present work are contained in the manuscript

## Declaration of competing interest

The authors declare that they have no known competing financial interests or personal relationships that could have appeared to influence the work reported in this paper.

## Ethics approval

This study entitled: “Evaluation of Pesticide Residues in Grain Sold at Selected Markets of Southwest Nigeria”, which was submitted for ethical approval on August 15, 2020 to the Research Ethics Committee of Kwara State University. For the development of this study, the fundamental ethical principles for research involving human beings, described and established by the Resolution 466/2012 and its complementary ones, was considered and approval was granted by the Kwara State University research ethics committee.

## Funding

Not Applicable (No funds were granted, received or used)

## Consent to participate

During the course of this research work, the participants were accorded the due respect so as to ensure co-operation and information collected were treated with utmost confidentiality. The cultures of the community were also respected during the course of the research work. Informed consent was obtained from all of the participants.

## Acknowledgements

We note with sadness the passing of our friend, colleague, and mentor **Prof. Mynepalli K. C. Sridhar**; working with him was wonderful and we strongly believe that he would have loved to coauthor this research paper – we miss him. We would like to say special thanks to all anonymous reviewer for providing very helpful comments for their valuable suggestions for improving this manuscript.

## Authorship contribution statement

**Modupe Abeke Oshatunberu:** Conceptualization, Investigation, Writing - original draft, Writing - review & editing. **Modupe Abeke Oshatunberu and Adebayo Oladimeji:** Investigation, Writing - original draft, Writing - review & editing. **Adebayo Oladimeji and Sawyerr Olawale Henry:** Resources, supervising, review & editing. **Modupe Abeke Oshatunberu, Morufu Olalekan Raimi:** Methodology, Project administration, Writing - original draft, Writing - review & editing. **Morufu Olalekan Raimi:** Data curation, Formal analysis, Writing - original draft, Writing - review & editing. **Modupe Abeke Oshatunberu, Adebayo Oladimeji, Sawyerr Olawale Henry:** Conceptualization, Data curation, Funding acquisition, Project administration, Writing - original draft, Writing - review & editing.

## Notes

### Competing Interest Statement

The authors have declared no competing interest.

### Funding Statement

This study did not receive any funding

### Author Declarations

This study entitled: Evaluation of Pesticide Residues in Grain Sold at Selected Markets of Southwest Nigeria, which was submitted for ethical approval on August 15, 2020 to the Research Ethics Committee of Kwara State University. For the development of this study, the fundamental ethical principles for research involving human beings, described and established by the Resolution 466/2012 and its complementary ones, was considered and approval was granted by the Kwara State University research ethics committee.

## References

1. Raimi MO, Abiola OS, Atoyebi B, Okon GO, Popoola AT Amuda-KA, Olakunle L Austin-AI & Mercy T. (2022). The Challenges and Conservation Strategies of Biodiversity: The Role of Government and Non-Governmental Organization for Action and Results on the Ground. In: Chibueze Izah, S. (eds) Biodiversity in Africa: Potentials, Threats, and Conservation. Sustainable Development and Biodiversity, vol 29. Springer, Singapore. https://doi.org/10.1007/978-981-19-3326-4_18.

2. Olalekan RM, Omidiji AO, Williams EA, Christianah MB, Modupe O (2019). The roles of all tiers of government and development partners in environmental conservation of natural resource: a case study in Nigeria. MOJ Ecology & Environmental Sciences 2019;4(3):114□121. DOI: 10.15406/mojes.2019.04.00142.

3. Raimi MO, Austin-AI, Olawale HS, Abiola OS, Abinotami WE, Ruth EE, Nimisingha DS & Walter BO (2022). Leaving No One Behind: Impact of Soil Pollution on Biodiversity in the Global South: A Global Call for Action. In: Chibueze Izah, S. (eds) Biodiversity in Africa: Potentials, Threats and Conservation. Sustainable Development and Biodiversity, vol 29. Springer, Singapore. https://doi.org/10.1007/978-981-19-3326-4_8.

4. Asiegbu OV, Ezekwe IC, Raimi MO (2022). Assessing pesticides residue in water and fish and its health implications in the Ivo River basin of South-eastern Nigeria. MOJ Public Health. 2022;11(4):136□142. DOI: 10.15406/mojph.2022.11.00390.

5. Olalekan MR, Abiola I, Ogah A, Dodeye EO (2021) Exploring How Human Activities Disturb the Balance of Biogeochemical Cycles: Evidence from the Carbon, Nitrogen and Hydrologic Cycles. Research on World Agricultural Economy. Volume 02, Issue 03. DOI: http://dx.doi.org/10.36956/rwae.v2i3.426. http://ojs.nassg.org/index.php/rwae.

6. Suleiman Romoke Monsurat, Raimi Morufu Olalekan and Sawyerr Henry Olawale (2019) A Deep Dive into the Review of National Environmental Standards and Regulations Enforcement Agency (NESREA) Act. International Research Journal of Applied Sciences. pISSN: 2663-5577, eISSN: 2663-5585. DOI No. Irjas.2019.123.123. https://www.scirange.com. https://scirange.com/abstract/irjas.2019.108.125

7. Okoyen E, Raimi M O, Omidiji A O, Ebuete A W (2020). Governing the Environmental Impact of Dredging: Consequences for Marine Biodiversity in the Niger Delta Region of Nigeria. Insights Mining Science and technology 2020; 2(3): 555586. DOI: 10.19080/IMST.2020.02.555586. https://juniperpublishers.com/imst/pdf/IMST.MS.ID.555586.pdf.

8. Lateefat MH, Opasola AO, Misbahu G, Morufu OR (2022) A Wake-Up Call: Determination of Antibiotics Residue Level in Raw Meat in Abattoir and Selected Slaughterhouses in Five Local Government in Kano State, Nigeria. bioRxiv 2022.01.04.474991; doi: https://doi.org/10.1101/2022.01.04.474991. Link: https://en.x-mol.com/paper/article/1479559526563860480

9. Lateefat MH, Opasola AO, Adiama BY, Ibrahim A, Morufu OR (2022) Elixirs of Life, threats to Human and Environmental Well-being: Assessment of Antibiotic Residues in Raw Meat Sold Within Central Market Kaduna Metropolis, Kaduna State, Nigeria. bioRxiv 2022.01.04.474997; doi: https://doi.org/10.1101/2022.01.04.474997.

10. Omotoso AJ, Omotoso EA, Morufu OR (2021) Potential Toxic levels of Cyanide and Heavy Metals in Cassava Flour Sold in Selected Markets in Oke Ogun Community, Oyo State, Nigeria, 01 July 2021, PREPRINT (Version 1) available at Research Square [https://doi.org/10.21203/rs.3.rs-658748/v1].

11. Adiama, YB., Sawyerr, OH., Olaniyi, OA., Fregene, AF., Alabede, M., & Raimi, MO. (2022). Assessment of Microbiological Quality of Ready to Eat Food Served in Ships Along Warri, Koko and Port Harcourt Water Ways, Nigeria. Online Journal of Microbiological Research, 1(1), 1–7. Retrieved from https://www.scipublications.com/journal/index.php/ojmr/article/view/230.

12. Lateefat HM, Faith A, Yusuf AB and Raimi OM (2022) Food for the Stomach Nourishing our Future: Assessment of Potassium Bromate in Local and Packaged Bread Sold in Ilorin Metropolis. Public H Open Acc, 6(1): DOI: 10.23880/phoa-16000197 Medwin Publishers ISSN: 2578-5001.

13. Habeeb ML, Opasola AO, Garba M, Olalekan MR. (2022). A Wake-Up Call: Determination of Antibiotics Residue Level in Raw Meat in Abattoir and Selected Slaughterhouses in Five Local Government in Kano State, Nigeria. J Vet Heal Sci, 3(1), 54–61.

14. Sawyerr O. H, Odipe O. E, Olalekan R. M, et al. (2018) Assessment of cyanide and some heavy metals concentration in consumable cassava flour “lafun” across Osogbo metropolis, Nigeria. MOJ Eco Environ Sci. 2018;3(6):369□372. DOI: 10.15406/mojes.2018.03.00115

15. Raimi MO, Ihuoma BA, Esther OU, Abdulraheem AF, Opufou T, Deinkuro NS, Adebayo PA and Adeniji AO (2020) “Health Impact Assessment: Expanding Public Policy Tools for Promoting Sustainable Development Goals (SDGs) in Nigeria”. EC Emergency Medicine and Critical Care 4.9 (2020).

16. Adedoyin OO, Olalekan RM, Olawale SH, et al (2020). A review of environmental, social and health impact assessment (Eshia) practice in Nigeria: a panacea for sustainable development and decision making. MOJ Public Health. 2020;9(3):81□87. DOI: 10.15406/mojph.2020.09.00328. https://medcraveonline.com/MOJPH/MOJPH-09-00328.pdf.

17. Olalekan RM, Oluwatoyin OA, Olawale SH, Emmanuel OO, Olalekan AZ (2020) A Critical Review of Health Impact Assessment: Towards Strengthening the Knowledge of Decision Makers Understand Sustainable Development Goals in the Twenty-First Century: Necessity Today; Essentiality Tomorrow. Research and Advances: Environmental Sciences. 2020(1): 72–84. DOI: 10.33513/RAES/2001-13. https://ospopac.com/journal/environmental-sciences/early-online.

18. Olalekan R. M, Oluwatoyin O and Olalekan A (2020) Health Impact Assessment: A tool to Advance the Knowledge of Policy Makers Understand Sustainable Development Goals: A Review. ES Journal of Public Health; 1(1); 1002. https://escientificlibrary.com/public-health/in-press.php.

19. Omidiji A.O and Raimi M.O (2019) Practitioners Perspective of Environmental, Social and Health Impact Assessment (ESHIA) Practice in Nigeria: A Vital Instrument for Sustainable Development. Paper Presented at the Association for Environmental Impact Assessment of Nigeria (AEIAN) On Impact Assessment: A Tool for Achieving the Sustainable Development Goals in Nigeria, 7th and 8th November, 2019 In University of Port Harcourt. https://aeian.org/wp-content/uploads/2019/08/EIA-Presentations-Portharcourt.pdf.

20. Raimi M.O, Omidiji A.O, Adio Z.O (2019) Health Impact Assessment: A Tool to Advance the Knowledge of Policy Makers Understand Sustainable Development Goals. Conference paper presented at the: Association for Environmental Impact Assessment of Nigeria (AEIAN) On Impact Assessment: A Tool for Achieving the Sustainable Development Goals in Nigeria, 7th and 8th November, 2019 in University of Port Harcourt. DOI: 10.13140/RG.2.2.35999.51366 https://www.researchgate.net/publication/337146101.

21. Hussain MI, Morufu OR, Henry OS (2021). Probabilistic Assessment of Self-Reported Symptoms on Farmers Health: A Case Study in Kano State for Kura Local Government Area of Nigeria. Environmental Analysis & Ecology Studies 9(1). EAES. 000701. 2021. DOI: 10.31031/EAES.2021.09.000701. Pp. 975–985. ISSN: 2578-0336.

22. Raimi Morufu Olalekan, Sawyerr Henry Olawale and Isah Hussain Muhammad (2020) Health risk exposure to cypermethrin: A case study of kano state, Nigeria. Journal of Agriculture. 7th International Conference on Public Healthcare and Epidemiology. September 14-15, 2020 | Tokyo, Japan.

23. Raimi Morufu Olalekan, and Sabinus Chibuzor Ezugwu. (2017) Influence of Organic Amendment on Microbial Activities and Growth of Pepper Cultured on Crude Oil Contaminated Niger Delta Soil. International Journal of Economy, Energy and Environment. Vol. 2, No. 4, 2017, pp. 56–76. DOI: 10.11648/j.ijeee.20170204.12.

24. Isah HM, Raimi MO, Sawyerr HO, Odipe OE, Bashir BG, Suleiman H (2020) Qualitative Adverse Health Experience Associated with Pesticides Usage among Farmers from Kura, Kano State, Nigeria. Merit Research Journal of Medicine and Medical Sciences (ISSN: 2354-323X) Vol. 8(8) pp. 432–447, August, 2020. DOI: 10.5281/zenodo.4008682. https://meritresearchjournals.org/mms/content/2020/August/Isah%20et%20al.htm.

25. Isah, H. M., Sawyerr, H. O., Raimi, M. O., Bashir, B. G., Haladu, S. & Odipe, O. E. (2020). Assessment of Commonly Used Pesticides and Frequency of Self-Reported Symptoms on Farmers Health in Kura, Kano State, Nigeria. Journal of Education and Learning Management (JELM), HolyKnight, vol. 1, 31–54. doi.org/10.46410/jelm.2020.1.1.05. https://holyknight.co.uk/journals/jelm-articles/.

26. Hussain MI, Morufu OR, Henry OS (2021) Patterns of Chemical Pesticide Use and Determinants of Self-Reported Symptoms on Farmers Health: A Case Study in Kano State for Kura Local Government Area of Nigeria. Research on Worl d Agri cul t ural Economy. Vol 2, No. 1. DOI: http://dx.doi.org/10.36956/rwae.v2i1.342. http://ojs.nassg.org/index.php/rwae/issue/view/31

27. Hussain MI, Morufu OR, Henry OS (2021) Probabilistic Assessment of Self-Reported Symptoms on Farmers Health: A Case Study in Kano State for Kura Local Government Area of Nigeria. Research on Worl d Agri cul t ural Economy. Vol 2, No. 1. DOI: http://dx.doi.org/10.36956/rwae.v2i1.336. http://ojs.nassg.org/index.php/rwae-cn/article/view/336/pdf.

28. Morufu OR, Tonye VO, Ogah A, Henry AE, Abinotami WE (2021) Articulating the effect of Pesticides Use and Sustainable Development Goals (SDGs): The Science of Improving Lives through Decision Impacts. Research on Worl d Agri cul t ural Economy. Vol 2, No. 1. DOI: http://dx.doi.org/10.36956/rwae.v2i1.347. http://ojs.nassg.org/index.php/rwae/issue/view/31

29. Morufu OR (2021). “Self-reported Symptoms on Farmers Health and Commonly Used Pesticides Related to Exposure in Kura, Kano State, Nigeria”. Annals of Community Medicine & Public Health. 1(1): 1002. http://www.remedypublications.com/open-access/self-reported-symptoms-on-farmers-health-and-commonly-used-pesticides-related-6595.pdf. http://www.remedypublications.com/annals-of-community-medicine-public-health-home.php.

30. Olalekan RM, Muhammad IH, Okoronkwo UL, Akopjubaro EH (2020). Assessment of safety practices and farmer’s behaviors adopted when handling pesticides in rural Kano state, Nigeria. Arts & Humanities Open Access Journal. 2020;4(5):191□201. DOI: 10.15406/ahoaj.2020.04.00170.

31. Erhunmwunse., A. Dirisu and J.O. Olomukoro (2012): Implication of pesticide usage in Nigeria. Tropical Fresh water Biology, 21 (1) 15–25.

32. Reagan Waskom, Troy Bauder, Robert Pearson, and Colorado Department, (2017): Best Management Practices. Agricultural Pesticide Use. May 2017 Bulletin #XCM-177

33. Morufu OR, Aziba-anyam GR and Teddy CA (2021) ‘Silent Pandemic’: Evidence-Based Environmental and Public Health Practices to Respond to the Covid-19 Crisis. IntechOpen. DOI: http://dx.doi.org/10.5772/intechopen.100204. ISBN 978-1-83969-144-7. https://www.intechopen.com/online-first/silent-pandemic-evidence-based-environmental-and-public-health-practices-to-respond-to-the-covid-19-Published: December 1st 2021; ISBN: 978-1-83969-144-7; Print ISBN: 978-1-83969-143-0; eBook (PDF) ISBN: 978-1-83969-145-4. Copyright year: 2021

34. Morufu OR, Aziba-anyam GR, Teddy CA (2021). Evidence-based Environmental and Public Health Practices to Respond to the COVID-19 Crisis, 07 May 2021, PREPRINT (Version 1) available at Research Square [https://doi.org/10.21203/rs.3.rs-504983/v1] https://europepmc.org/article/PPRID/PPR335534; EMSID:EMS123969.

35. Raimi, MO., Mcfubara, KG., Abisoye, OS., Ifeanyichukwu EC., Henry SO., & Raimi, GA (2021) Responding to the call through Translating Science into Impact: Building an Evidence-Based Approaches to Effectively Curb Public Health Emergencies [COVID-19 Crisis]. Global Journal of Epidemiology and Infectious Disease, 1(1). DOI: 10.31586/gjeid.2021.010102. Retrieved from https://www.scipublications.com/journal/index.php/gjeid/article/view/72.

36. Raimi MO & Raimi AG (2020). The Toughest Triage in Decision Impacts: Rethinking Scientific Evidence for Environmental and Human Health Action in the Times of Concomitant Global Crises. CPQ Medicine, 11(1), 01–05.

37. Raimi MO, Moses T, Okoyen E, Sawyerr HO, Joseph BO, Oyinlola BO (2020) “A Beacon for Dark Times: Rethinking Scientific Evidence for Environmental and Public Health Action in the Coronavirus Diseases 2019 Era” Medical and Research Microbiology, Vol. 1, Issues 3.

38. Ajayi FA, Raimi MO, Steve-Awogbami OC, Adeniji AO, Adebayo PA (2020) Policy Responses to Addressing the Issues of Environmental Health Impacts of Charcoal Factory in Nigeria: Necessity Today; Essentiality Tomorrow. Communication, Society and Media. Vol 3, No 3. DOI: https://doi.org/10.22158/csm.v3n3p1. http://www.scholink.org/ojs/index.php/csm/article/view/2940.

39. Asogwa E.U. and Dongo L.N (2009) Problems associated with pesticide usage and application in Nigerian cocoa production: A review. Afric J of Agric research. vol. 4 (8), pp675–683

40. Adedoyin O.T., Ojuawo A., Adesiyun O.O., Mark F., Anigilaje E.A (2008): Poisoning due to yam flour consumption in five families in Ilorin, central Nigeria. West Afr. Med. J.; 27:41–43.

41. Adeleke S.I. (2009): Food poisoning due to yam flour consumption in Kano (Northwest) Nigeria Online J of Health and Allied Sci 8 (2).

42. Fatiregun A.A., Oyebade O.A., Oladokun L. (2010): Investigation of an outbreak of food poisoning in a resource-limited setting. Trop. J. Health Sci. 17:1117–4153.

43. Ilesanmi, F. F., Ilesanmi, O. S., & Afolabi, A. A. (2021). The effects of the COVID-19 pandemic on food losses in the agricultural value chains in Africa: The Nigerian case study. Public Health in Practice, 2, 100087. https://doi.org/10.1016/j.puhip.2021.100087.

44. van Bueren, E. T. L., Struik, P. C., van Eekeren, N., & Nuijten, E. (2018). Towards resilience through systems-based plant breeding. A review. Agronomy for Sustainable Development, 38, 42.

45. Verneau, F., Amato, M., & La Barbera, F. (2021). Edible insects and global food security. Insects, 12(5), 10–12. https://doi.org/10.3390/insects12050472.

46. Owoo, N. S. (2018). Food insecurity and family structure in Nigeria. SSM - Population Health, 4, 117–125. https://doi.org/10.1016/j.ssmph.2017.12.004.

47. Fasanya, I. O., & Odudu, T. F. (2020). Modeling return and volatility spillovers among food prices in Nigeria. Journal of Agriculture and Food Research, 2, 100029. https://doi.org/10.1016/j.jafr.2020.100029.

48. Obinna-Echem, P. C., Beal, J., & Kuri, V. (2015). Effect of processing method on the mineral content of Nigerian fermented maize infant complementary food - Akamu. LWT - Food Science and Technology, 61, 145–151. https://doi.org/10.1016/j.lwt.2014.11.001.

49. Sosan, M.B., Oyekunle, J. A.., & Odewale, G.. (2018). Occurrence and levels of organochlorine pesticide residues in maize samples from open markets and stores in Ile-Ife and Ondo, Southwestern Nigeria. Nigeria Journal of Entomology, 34(1), 25–37. https://doi.org/10.36108/nje/8102/43(0140).

50. Moussa, H., Kindomihou, V., Houehanou, T. D., Chaibou, M., Souleymane, O., Soumana, I., Dossou, J., & Sinsin, B. (2021). Farmers’ perceptions of fodder performances of pearl millet (Pennisetum glaucum (L.) R. Br) accessions in Niger. Heliyon, 7(9), e07965. https://doi.org/10.1016/j.heliyon.2021.e07965.

51. Salihu, I., Kasum, L. A., & Agbara, I. G. (2021). Production and evaluation of the quality of pearl millet-based fura (A northern Nigerian cereal-based Spiced Steamed Dough) as Affected by bambara groundnut flour supplementation. Asian Journal of Applied Science and Technology, 05(03), 13–31. https://doi.org/10.38177/ajast.2021.5302.

52. Kulkarni, D. B., Sakhale, B. K., & Chavan, R. F. (2021). Studies on development of low gluten cookies from pearl millet and wheat flour. Food Research, 5(4), 114–119. https://doi.org/10.26656/fr.2017.5(4).028.

53. Onyeneke, R. U., Amadi, M. U., Njoku, C. L., & Osuji, E. E. (2021). Climate change perception and uptake of climate-Smart agriculture in rice production in Ebonyi State, Nigeria. Amosphere, 12, 1503.

54. Lindgren, E., Harris, F., Dangour, A. D., Gasparatos, A., Hiramatsu, M., Javadi, F., Loken, B., Murakami, T., Scheelbeek, P., & Haines, A. (2018). Sustainable food systems—a health perspective. Sustainability Science. https://doi.org/10.1007/s11625-018-0586-x.

55. Mobolade, A. J., Bunindro, N., Sahoo, D., & Rajashekar, Y. (2019). Traditional methods of food grains preservation and storage in Nigeria and India. Annals of Agricultural Sciences, 64(2), 196–205. https://doi.org/10.1016/j.aoas.2019.12.003.

56. Planas, S., Román, C., Sanz, R., & Rosell-Polo, J. R. (2022). Bases for pesticide dose expression and adjustment in 3D crops and comparison of decision support systems. Science of the Total Environment, 806, 150357. https://doi.org/10.1016/j.scitotenv.2021.150357.

57. You, L., Zheng, F., Su, C., Wang, L., Li, X., Chen, Q., Kou, J., Wang, X., Wang, Y., Wang, Y., Mei, S., Zhang, B., Liu, X., & Xu, G. (2022). Metabolome-wide association study of serum exogenous chemical residues in a cohort with 5 major chronic diseases. Environment International, 158, 106919. https://doi.org/10.1016/j.envint.2021.106919

58. Sang, C., Yu, Z., An, W., Borgen Sørensen, P., Jin, F., & Yang, M. (2022). Development of a data driven model to screen the priority control pesticides in drinking water based on health risk ranking and contribution rates. Environment International, 158, 106901. https://doi.org/10.1016/j.envint.2021.106901.

59. FAOSTAT (2020) Statistical Database. Food and Agriculture Organization of the United Nations, Rome.

60. Čuš, F., Česnik, H. B., & Bolta, Š. V. (2022). Pesticide residues, copper and biogenic amines in conventional and organic wines. Food Control, 132, 108534. https://doi.org/10.1016/j.foodcont.2021.108534.

61. Al-Mrammathi, M. M. A. F., & Al-Hasnawi, M. R. A. (2021). Insects and insect pest management system. IOP Conference Series: Earth and Environmental Science, 735, 012028. https://doi.org/10.1088/1755-1315/735/1/012028.

62. Grung, M., Lin, Y., Zhang, H., Steen, A. O., Huang, J., Zhang, G., & Larssen, T. (2015). Pesticide levels and environmental risk in aquatic environments in China - A review. Environment International, 81, 87– 97. https://doi.org/10.1016/j.envint.2015.04.013.

63. Halwachs, S., Schäfer, I., Kneuer, C., Seibel, P., & Honscha, W. (2016). Assessment of ABCG2-mediated transport of pesticides across the rabbit placenta barrier using a novel MDCKII in vitro model. Toxicology and Applied Pharmacology, 305, 66–74. https://doi.org/10.1016/j.taap.2016.06.007.

64. Rial-Berriel, C., Acosta-Dacal, A., Zumbado, M., Henríquez-Hernández, L. A., Rodríguez-Hernández, Á., Macías-Montes, A., Boada, L. D., Travieso-Aja, M. del M., Martin Cruz, B., & Luzardo, O. P. (2021). A method scope extension for the simultaneous analysis of pops, current-use and banned pesticides, rodenticides, and pharmaceuticals in liver. Application to food safety and biomonitoring. In Toxics (Vol. 9, Issue 10). https://doi.org/10.3390/toxics9100238.

65. Dassanayake, M. K., Chong, C. H., Khoo, T. J., Figiel, A., Szumny, A., & Choo, C. M. (2021). Synergistic field crop pest management properties of plant-derived essential oils in combination with synthetic pesticides and bioactive molecules: A review. Foods, 10, 2016. https://doi.org/10.3390/foods10092016.

66. Akinneye, J. O., Adedolapo, A., & Adesina Femi, P. (2018). Quantification of Organophosphate and Carbamate residue on stored grains in Ondo State, Nigeria. Journal of Biology and Medicine, 2(1), 001–006. https://doi.org/10.17352/jbm.000003.

67. Li, A. J., & Kannan, K. (2018). Urinary concentrations and profiles of organophosphate and pyrethroid pesticide metabolites and phenoxyacid herbicides in populations in eight countries. Environment International, 121, 1148–1154. https://doi.org/10.1016/j.envint.2018.10.033.

68. Olisah, C., Okoh, O. O., & Okoh, A. I. (2020). Occurrence of organochlorine pesticide residues in biological and environmental matrices in Africa: A two-decade review. Heliyon, 6(3), e03518. https://doi.org/10.1016/j.heliyon.2020.e03518.

69. Krief, A. (2021). Pyrethroid insecticides. Arkivoc, 1, 48–54. https://doi.org/10.1039/b202996k

70. Fatunsin, O. T., Oyeyiola, A. O., Moshood, M. O., Akanbi, L. M., & Fadahunsi, D. E. (2020). Dietary risk assessment of organophosphate and carbamate pesticide residues in commonly eaten food crops. Scientific African, 8, e00442. https://doi.org/10.1016/j.sciaf.2020.e00442.

71. Bamidele, A., Abayomi, A., Iyabo, A., & Giwa, M. (2019). Parasitic fauna, histopathological alterations, and organochlorine pesticides contamination in Chrysichthys nigrodigitatus (Lacepede, 1803) (Bagridae) from Lagos, Lagoon, Nigeria. Scientific African, 5, e00130. https://doi.org/10.1016/j.sciaf.2019.e00130.

72. Ingenbleek, L., Hu, R., Pereira, L. L., Paineau, A., Colet, I., Koné, A. Z., Adegboye, A., Hossou, S. E., Dembélé, Y., Oyedele, A. D., Kisito, C. S. K. J., Eyangoh, S., Verger, P., Leblanc, J. C., & Le Bizec, B. (2019). Sub-Saharan Africa total diet study in Benin, Cameroon, Mali and Nigeria: Pesticides occurrence in foods. Food Chemistry: X, 2, 100034. https://doi.org/10.1016/j.fochx.2019.100034.

73. Sosan, Mosudi B., Adeleye, A. O., Oyekunle, J. A. O., Udah, O., Oloruntunbi, P. M., Daramola, M. O., & Saka, W. T. (2020). Dietary risk assessment of organochlorine pesticide residues in maize-based complementary breakfast food products in Nigeria. Heliyon, 6(12), e05803. https://doi.org/10.1016/j.heliyon.2020.e05803.

74. AOAC. (2000). Official Methods of Analysis. Washington DC: Association of Official Analytical Chemists.

75. European Committee for Standardization. (2018). ILNAS-EN 15662 Foods of Plant Origin - Multimethod for the Determination of Pesticide Residues Using GC- and LC-based Analysis Following Acetonitrile extraction/partitioning and Clean-up by Dispersive SPE - Modular QuEChERS-method. https://ilnas.services-publics.lu/ecnor/downloadP.

76. Gesesew, H. A., Woldemichael, K., Massa, D., & Mwanri, L. (2016). Farmers knowledge, attitudes, practices and health problems associated with pesticide use in rural irrigation villages, Southwest Ethiopia. PLoS ONE, 11(9), 1–13. https://doi.org/10.1371/journal.pone.0162527.

77. Yusuf I.B., Azeez I.A, and Bolaji O.M (2019) Assessment of Knowledge and Practice of Pesticide Application Among Beans and Maize Sellers in an Urban Southwestern Nigerian Market. Afr. J. Biomed. Res. Vol. 22 (January 2019); 27–32.

78. Alex, A. A., Longinus, N. K., Olatunde, A. M., & Chinedu, N. V. (2018). Pesticides related knowledge, attitude, and safety practices among small-scale vegetable farmers in lagoon wetlands, Lagos, Nigeria. Journal of Agriculture and Environment for International Development, 112(1), 81–99. https://doi.org/10.12895/jaeid.20181.697.

79. Mekonnen Y and Agonafir T (2002) Pesticide sprayers’ knowledge, attitude, and practice of pesticide use on agricultural farms of Ethiopia. Occup. Med. Vol. 52 No. 6, pp. 311–315.

80. Tariq, M.I.; Afzal, S.; Hussain, I.; Sultana, N (2007). Pesticides exposure in Pakistan: A review. Environ. Int., 33, 1107–1122.

81. Mekonnen, Y., & Ejigu, D. (2005). Plasma cholinesterase level of Ethiopian farm workers exposed to chemical pesticide. Occupational Medicine, 55, 504505.

82. Tariq, M.I., Hussain, I., & Afzal, S. (2003). Policy measures for the management of water pollution in Pakistan. Pakistan Journal of Environmental Science, 3, 11–51.

83. Huang, Y., Liu, L., & Pei, E. (2008). Vegetative pesticide residue and application behaviour in Beijing. Chinese Journal of Food Hygiene, 20, 319321.

84. Xu, Y. (2004). Survey of improper pesticide application in rice production. Jiangxi Agricultural Technology, 11, 27–28.

85. Agarwal, P.K., & Pandey, D.I.V.Y.A. (2017). Impact of Pesticide: An Overview. Trends in Biosci, 10(6), 1341–1344.

86. Abhilash, P.C., & Singh, N. (2009). Pesticide use and application: An Indian scenario. Journal of Hazardous Materials, 165(1:3), 1–12.

87. Li, M., Li, X., & Fu, X. (2008). A study of farmers’ behaviour, knowledge and attitude toward pesticide application in Chengdu region. Journal of Preventive Medicine Information, 24, 521–525.

88. Dongmei, Z. (2006). Development of bio-pesticide industry in China. Unpublished PhD thesis, Fujian University of Agriculture and Forestry, Fujian, China.

89. Zhongze, F., & Qingjiang, L. (2007) Farmers safety awareness and influence factors of agricultural quality. Economic Problems of Agriculture, 4, 2225.

90. Yang, P., Li, P., & Zhou, J. (2007). Vegetable cultivation habits of small-scale farms in Yunnan Province and pest management behaviour. Plant Protection, 6, 9499.

91. Zhou, B. (2007) Study of rice farmers pesticide application behaviour and attitude a case study based on surveys of rice farmers in Taizhou. Unpublished master’s thesis, Yangzhou University, Jiangsu Province, China.

92. Reffstrup, T. K., Larsen, J. C., & Meyer, O. (2010). Risk assessment of mixtures of pesticides. Current approaches and future strategies. Regulatory Toxicology and Pharmacology, 56, 174–192. https://doi.org/10.1016/j.yrtph.2009.09.013.

93. World Health Organization/Food and Agriculture Organization (WHO/FAO) (2010). Pesticide residues in food: maximum residue limits, extraneous maximum residue limits. Rome, Food and Agriculture Organization of the United Nations

94. Shem OW (2001). Use and distribution of organochlorines pesticides. The future in Africa. Pure and Applied Chemistry, 73(7):1147–1155.

95. Jallow, M. F. A., Awadh, D. G., Albaho, M. S., Devi, V. Y., & Thomas, B. M. (2017). Pesticide knowledge and safety practices among farm workers in Kuwait: Results of a survey. International Journal of Environmental Research and Public Health, 14(4). https://doi.org/10.3390/ijerph14040340.

96. Nkemleke, E.E. (2020) Assessing Small-Scale Farmers’ Attitudes, Practices and Vulnerability to Pesticides Use in Market Gardening Crops in M’muockngie (Southwestern Cameroon). Journal of Advances in Education and Philosophy, 4(6), 295–305. https://doi.org/10.36348/jaep.2020.v04i06.010.

97. Macfarlane E., Chapman A., Benke B., Meaklim J., Sim M., McNeil J (2008). Training and other predictors of personal protective equipment use in Australian grain farmers using pesticides. Occup Environ Med;65(2): 141–146.

98. Aminu, F.O.1 and Edun, T.A. (2019) Environmental Effect of Pesticide Use by Cocoa Farmers in Nigeria. Journal of Research in Forestry, Wildlife & Environment Vol. 11(4).http://www.ajol.info/index.php/jrfwe. ISBN: 2141 – 1778

99. Handford, C.E., Elliott, C.T., Campbell, K (2015). A review of the global pesticide legislation and the scale of challenge in reaching the global harmonization of food safety standards. Integr Environ Assess Manag 11(4): 525–536.

100. Saath, K.C.D. O., & Fachinello, A.L (2018) Crescimento da demanda mundial de alimentos e restrições do fator terra no Brasil. Revista de Economia e Sociologia Rural, 56(2), 195–212. https://doi.org/10.1590/1234-56781806-94790560201.

101. Van Maele-Fabry G, Lantin AC, Hoet P, Lison D (2011). Residential exposure to pesticides and childhood leukaemia: a systematic review and meta-analysis. Environ Int.Jan;37(1):280–91. doi: 10.1016/j.envint.2010.08.016. PMID: 20889210.

102. Oguntade Adesola Saheed, Sawyerr Henry Olawale, Salami John Tolulope (2020) Assessment of Pesticide Residues in Grains Sold in Major Commercial Markets in Ilorin, Kwara State, Nigeria. International Journal of Research and Innovation in Applied Science (IJRIAS) | Volume V, Issue IX, September 2020|ISSN 2454-6194.

